# A systematic review of enteric pathogens and antibiotic resistance genes in outdoor urban aerosols

**DOI:** 10.1101/2021.10.26.21265483

**Authors:** Olivia Ginn, Sarah Lowry, Joe Brown

**Affiliations:** Department of Civil & Environmental Engineering & Earth Science, University of Notre Dame, 156 Fitzpatrick Hall, Notre Dame, IN 46556, USA; Department of Civil and Environmental Engineering, Stanford University, Stanford, California, United States; Deparment of Environmental Sciences and Engineering, Gillings School of Global Public Health, University of North Carolina, Chapel Hill, North Carolina, 27599, United States

**Author notes:** Correspondence to: (Primary) Dr. Joe Brown, Department of Environmental Sciences and Engineering, University of North Carolina, 135 Dauer Drive, Chapel Hill, NC, 27599, USA. (Secondary) Dr. Olivia Ginn, Department of Civil & Environmental Engineering & Earth Science, University of Notre Dame, 156 Fitzpatrick Hall, Notre Dame, IN, 46556, USA.

**Keywords:** enteric pathogens, antibiotic resistance, bioaerosols, urban air, public health, sanitation

## Abstract

Aerosol transport of enteric microbiota including fecal pathogens and antimicrobial resistance genes (ARGs) has been documented in a range of settings but remains poorly understood outside indoor environments. We conducted a systematic review of the peer-reviewed literature to summarize evidence on specific enteric microbiota including enteric pathogens and ARGs that have been measured in aerosol samples in urban settings where the risks of outdoor exposure and antibiotic resistance (AR) spread may be highest. Following PRISMA guidelines, we conducted a key word search for articles published within the years 1990-2020 using relevant data sources. Two authors independently conducted the keyword searches of databases and conducted primary and secondary screenings before merging results. To be included, studies contained extractable data on enteric microbes and AR in outdoor aerosols regardless of source confirmation and reported on qualitative, quantitative, or viability data on enteric microbes or AR. Qualitative analyses and metric summaries revealed that enteric microbes and AR have been consistently reported in outdoor aerosols, generally via relative abundance measures, though gaps remain preventing full understanding of the role of the aeromicrobiological pathway in the fate and transport of enteric associated outdoor aerosols. We identified remaining gaps in the evidence base including a need for broad characterization of enteric pathogens in bioaerosols beyond bacterial genera, a need for greater sampling in locations of high enteric disease risk, and a need for quantitative estimation of microbial and nucleic acid densities that may be applied to fate and transport models and in quantitative microbial risk assessment.

**FUNDING:** This study was funded by the National Science Foundation under grant number 1653226. This funding source had no role in the design of this study and had no role during its execution, analyses, interpretation of the data, or decision to submit results.

## 1.0 INTRODUCTION

Studies in high-risk outdoor settings in the USA and in other high-income countries have revealed that bioaerosols containing enteric (intestinal) microbes are common where concentrated human or animal fecal waste and one or more mechanisms for aerosolization co-occur. Presence of enteric microbes in aerosols has been best characterized in ambient air surrounding specific point and area sources such as wastewater treatment plants^1–12^ land applied biosolids,^13–23^ composting facilities,^24,25^ meat markets,^26^ urban areas,^9,27,28^ landfills,^29,30^ and concentrated animal feeding operations.^31–33^ In many of these cases, adequate waste disposal technology exists but bioaerosols containing these targets are consistently detected. A growing body of literature acknowledges the potential risk accompanying enteric microbes and antibiotic resistant (AR) enteric microbes in aerosols where a source is suspected or confirmed. Studies of exposure to these point and area sources typically assess nearby residents^20,34^ or facility workers^35,36^ who might be at greatest risk.

Settings less well characterized in the context of enteric pathogens and associated AR are ambient urban outdoor environments where sources may be diffuse and related to generally inadequate or poorly performing environmental controls or waste management^37,38^. Establishing and maintaining effective and sustainable infrastructure to improve population health in cities is becoming increasingly difficult due to factors including accelerating urbanization and densification, climate change and its effects on infrastructure, and financing constraints; these and other limiting factors are most acute in low- and middle-income countries (LMICs). Information on ambient fluxes of enteric pathogens and AR in cities lacking good waste management is needed, targeting settings with potentially elevated exposure risks and AR spread. Cities in high-income countries (HICs) are not generally considered high risk for exposure to enteric pathogens via sanitary deficits: human excreta is typically directed into covered or underground wastewater infrastructure, creating a separation between humans and their waste to interrupt transmission of enteric infections via well understood pathways.^39^ However, in LMICs where wastewater and biosolids present greater risks due to the high number of circulating infectious diseases – and poorly controlled antibiotic stewardship^40^ – in the population contributing waste and the presence of unsequestered waste in closer proximity to more people, risk may be higher for exposure through inhalation or ingestion, including indirectly following surface deposition of bioaerosols.^41–44^ Recent studies have described novel detections along with aeromicrobiological transport of enteric pathogens in aerosols, drawing attention to this pathway in the context of disease transmission^44,45^ and AR emergence and dissemination. In order to advance understanding of the current and potential impact of enteric pathogens and AR in bioaerosols in high-risk settings, we seek to describe the rich emerging literature on this topic and identify gaps in evidence. Therefore, we conducted a systematic review of published estimates reporting detections of enteric microbiota and ARs in cities.

## 2.0 METHODS

We followed PRISMA guidelines for reporting of systematic reviews^46^ as recommended in the WHO Handbook for Guideline Development,^47^ including all meta-data from searches per the recommended PRISMA flow diagram. We pre-specified our methods (https://osf.io/6Q7E9/) and provide a full list of preferred reporting items in Supplemental Material. Our outcomes included any report of enteric microbe (bacteria, virus, protozoa) or any ARG from pre-determined lists meeting our site criteria, from either qualitative or quantitative studies assessing their presence. Site criteria were: urban areas with either no known source or a point or area source related to concentrated animal feeding operations (CAFOs), wastewater treatment plants (WWTPs), composting facilities, landfills or other identified source.

### 2.1 Database sources and search string strategy

*A priori*, we chose a list of relevant and well-known databases in which to conduct our search for academic peer-reviewed literature including: ISI Web of Science, PubMed, SCOPUS, EBSCOhost, and Science.gov. We conducted additional searches in ProQuest Dissertations and Theses Global.

We compiled the string of keywords for searching the literature databases from our own knowledge of the topic, and from suggestions by experts and existing reviews in related topics.^48–52^ These search terms were refined based on synonyms, abbreviations, and alternate spellings and were expanded and adjusted as initial searches were performed. Searches were in the form of a one line string that included all fields from the scope (e.g., aerosol, extramural, urban…), pathogens (e.g., enteric, fecal indicator, adenovirus…), exposures (e.g., inhalation, ingestion, viable…), mechanisms (e.g., aerosolization, bubbles, aeration…), sampling methods (e.g., impaction, impinger, filter…), AR associated (e.g., *intI1, tetA, qnrB*…), and antibiotics (e.g., tetracycline, fluoroquinolones, multidrug resistance…). Search strings used “or” and “and” statements to include multiple fields such as “fecal indicator bacteria OR FIB” AND “bioaerosol OR ambient OR air OR aerosol OR airborne”. The full search string is detailed in Supplemental Material.

Two authors independently conducted the keyword searches in each database and compiled findings in Zotero (Version 5·0·96). After deduplication of search results, the authors each conducted an initial screening based on titles and abstracts of papers. Papers passed the initial screening if the title or abstract suggested relevance to the keywords in the search. The full text of papers that passed initial screening were reviewed independently by the two authors if papers were: (1) published in English; (2) published from the 1990-2020; and (3) published or accepted for publication in a peer-reviewed journal, refereed conference proceedings, government or multilateral document, thesis or dissertation, book, or other document appearing on indexed databases we identified; (4) contained extractable data on enteric microbes including pathogens and AR in outdoor bioaerosols; (5) contained data on targets that could be related to no source, a specific point source, or an area source but must have been from an urban site; and (6) reported on qualitative (presence/absence), quantitative (culture or PCR-based), or viability (culture) data that was sufficient for interpretation within the three aforementioned groups.

After independent searches, we consulted and compared our findings to achieve consensus on inclusion. For any dissent on inclusion of a paper, a third party was consulted for discussion before again reaching a consensus decision on inclusion. Backward searches examined the references, authors and contributors, and keywords identified in the literature resulting from the initial keyword searches. Forward searches examined articles not initially identified that have referenced literature resulting from the initial keyword searches.

### 2.2 Data collection and analysis

For each study, data were extracted and recorded; the full compilation is included in Supplemental Material. We recorded document identifiers such as authors, publication year, document type, and whether the document was found through initial, backward, or forward searches. We categorized documents meeting inclusion criteria first by subject, either AR or enteric microbes detection. Some publications reported both. We then categorized these by analysis method: culture, PCR, or sequencing.

We extracted and recorded general study characteristics such as location, sampling method, sample size, and intended targets. Depending on the category, we recorded qualitative, quantitative, and semi-quantitative data, indicating relevant targets and results within the scope of our review. A formal meta-analysis of the disparate findings was not deemed possible given the limited evidence on this topic and the wide diversity of methods employed in measurement in this emerging area. Therefore, we synthesized review data through categorization and descriptive summary, a common approach when quantitative meta-analysis is not possible,^53^ revealing both a broad description of what is known as well as identifying gaps requiring further evidence.

## 3.0 RESULTS

### 3.1 Search results

Searches returned 1226 publications from 6 databases. Upon removal of duplicates, initial screening of the title and abstract, and full text screening, 52 publications met search criteria. Forward and backward searches yielded additional publications for a total of 101 to be included in this review.

### 3.2 Study characteristics

Of the 101 studies we included, ten reported multiple data types or multiple subjects and methodologies, reflected in Figure 1 and Table 1. Table 1 details the intended targets for each study. The full table listing each publication by first author and year can be found in Supplemental Material. Overall, 28% of the studies identified in this systematic review reported on AR related data and 82% reported on enteric microbe related data (1% reported both). We observed sequencing to be the most frequent analysis method, accounting for 75% across all studies. A majority of studies took place in upper-middle and high-income countries (n=89), with only ten studies based in LMICs including India, Mali, Bolivia, and Vietnam (Figure 2). A few studies collected data from both income categories. China accounted for almost 50% of all studies. Only 9% of studies were published since 2010.

**Figure 1.**
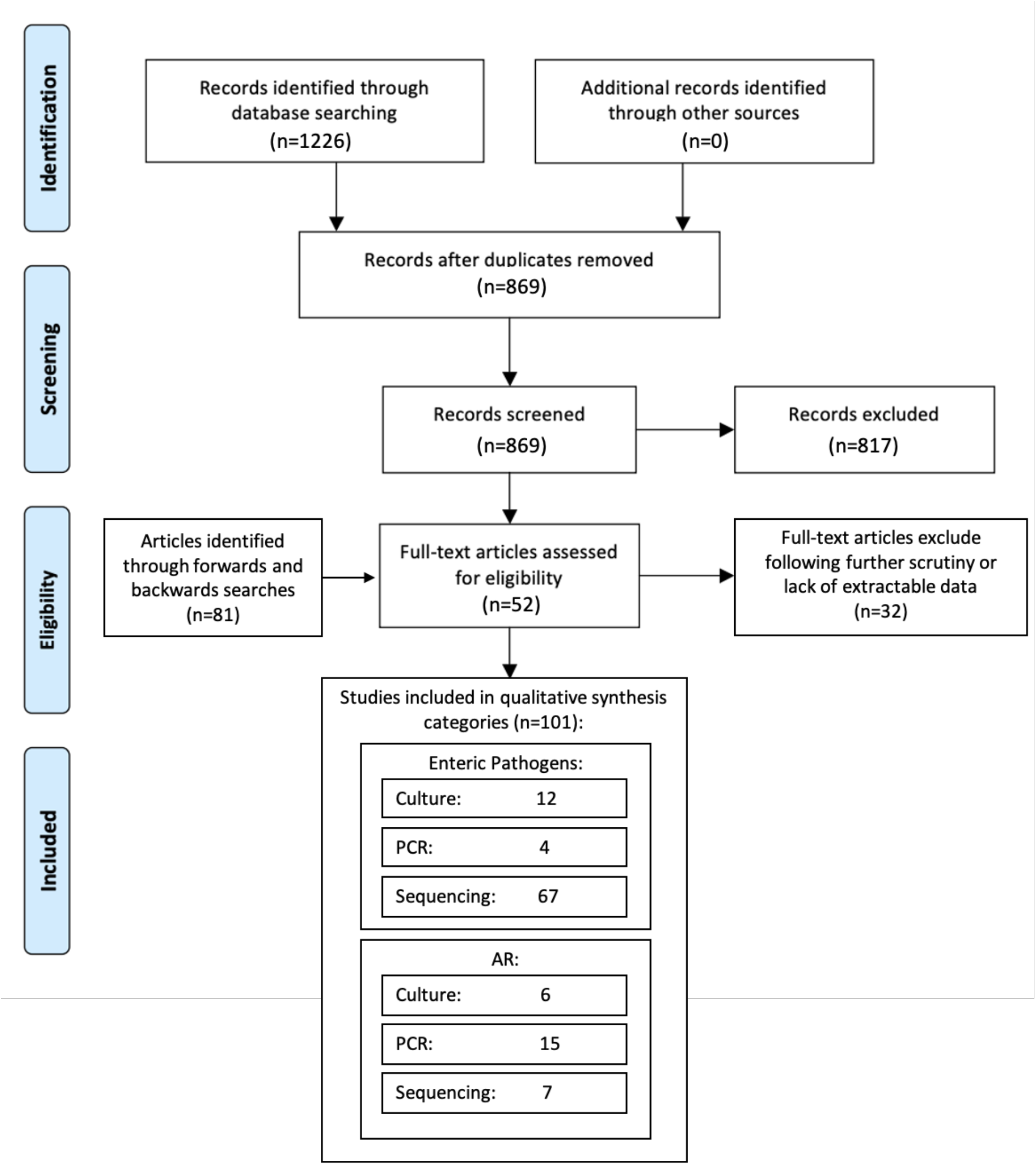
Modified PRISMA flow diagram applied to our systematic review process.

**Table 1:**
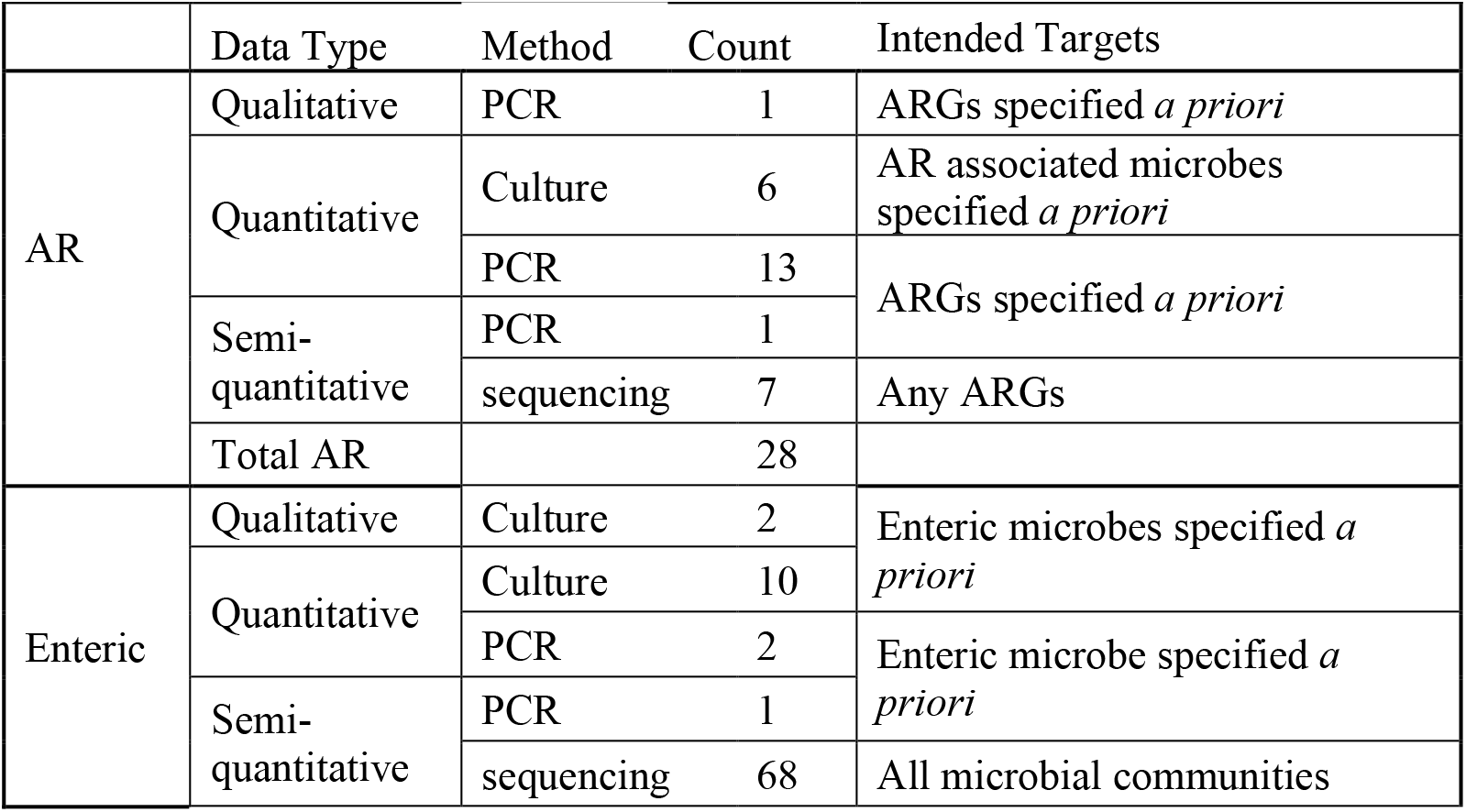
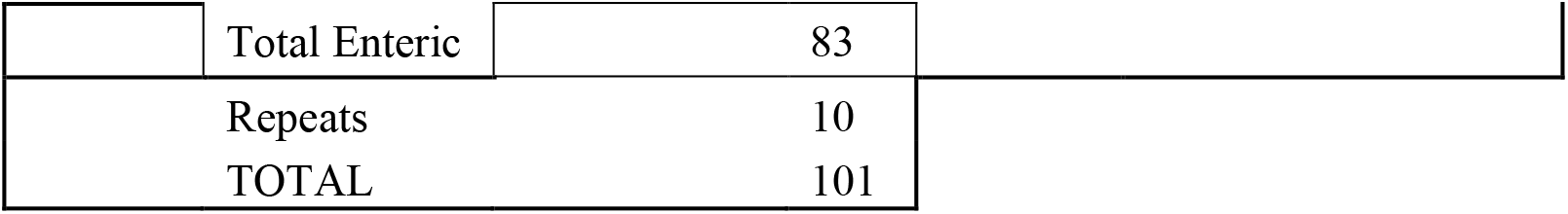
Studies included in the review by categories of target type (antibiotic resistance related or enteric microbe related) and by data type and analysis method. The intended targets are also categorized.

**Figure 2.**
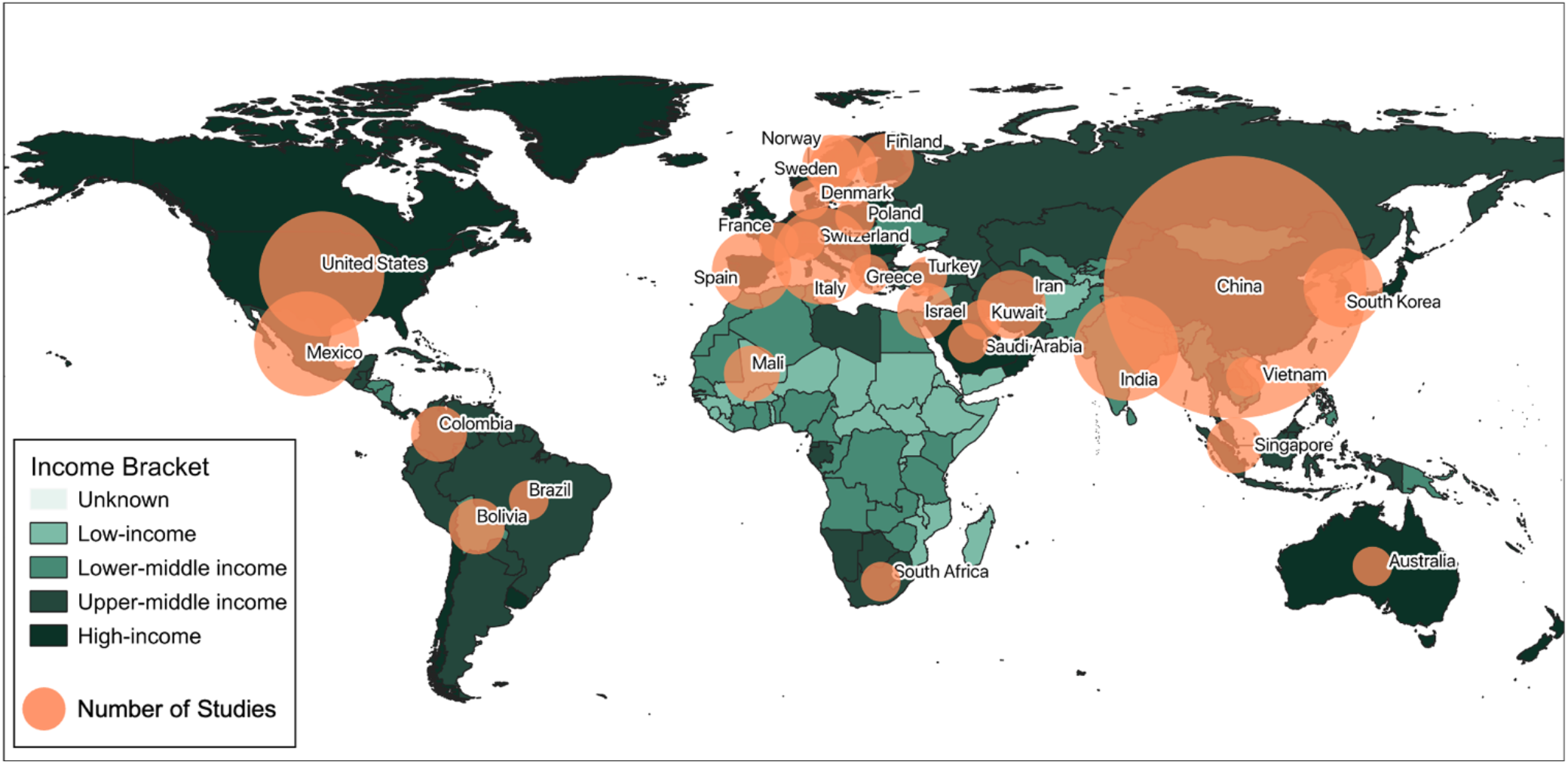
Total number of studies by country of measurement, with circle size proportional to number of studies (n =1 to 44 in China).

Bioaerosol sampling can typically be categorized by three different methods: filtration, impaction, and impingement. In our review, the majority of studies employed filtration to collect bioaerosol samples (n=60 studies). Filtration was followed by active impaction (n=28), impingement (n=10), and passive impaction via gravitational settling (n=8). Some studies used multiple methods, often when performing culture-dependent analyses in addition to culture-independent analyses.

### 3.3 Results from specific studies

#### Enteric microbes

Overall, studies reporting on enteric microbe detection in bioaerosols in urban settings targeted a limited range of microbes, focusing on well-established fecal indicators and in the case of detection via culture, easily culturable bacteria such as *Enterococcus, Enterobacter* and *Escherichia* genera.^54^ One study reported detection through culture of *Yersinia enterocolitica, Salmonella* spp. and others in Chinese live poultry markets in urban Tai’an,^55^ though in general the city includes extensive sanitation infrastructure throughout.^56^ For quantitative PCR (qPCR), reported enteric microbes included pathogens as well as possibly commensal enteric bacteria (*Escherichia coli, Enterococcus* spp., *Enterobacter* spp., *Yersinia enterocolitica*, and *Salmonella* spp.)^54,57^ and enteric virus (human adenovirus, human enterovirus, and human rotavirus).^58–60^

A majority of studies assessed overall microbial communities through sequencing, typically employing 16S rRNA. Of these 67 studies, nine classified the population taxonomy to order level,^61–68^ eight classified to family level,^69–74^ 37 classified to genus,^51,64,75–109^ 12 classified to species^28,54,59,83,98,110–118^ and only one identified a strain-specific^119^ species (Figure 3). When classified to order or family, *Enterobacteriales* and *Enterobacteriaceae* (respectively) were detected as less than 1% of the bacterial community. However, one study detected the families *Campylobacteraceae* (*Arcobacter*) and *Helicobacteraceae*^70^ and another study detected the orders *Clostridiales* and *Campylobacterales*.^65^ *Escherichia* or *Escherichia-Shigella, Enterococcus and Clostridium* were detected most frequently at the genus level. *Vibrio, Campylobacter, Yersinia, Klebsiella, and Aeromonas* were also detected in a few studies at the genus level. Enteric microbes detected at the genus or species level through sequencing included *Aeromonas* spp., *Enterobacter* spp., *Escherichia coli, Enterococcus faecium, Clostridium* spp., *Vibrio parahaemolyticus, Salmonella enterica, Campylobacter coli, Yersinia enterocolitica, Shigella* spp., *Klebsiella pneumonia*, and *Escherichia fergusonii*. One study in a tropical urban setting identified, through sequencing, the pathogenic strain of *Escherichia coli* O157:H7.^119^

**Figure 3.**
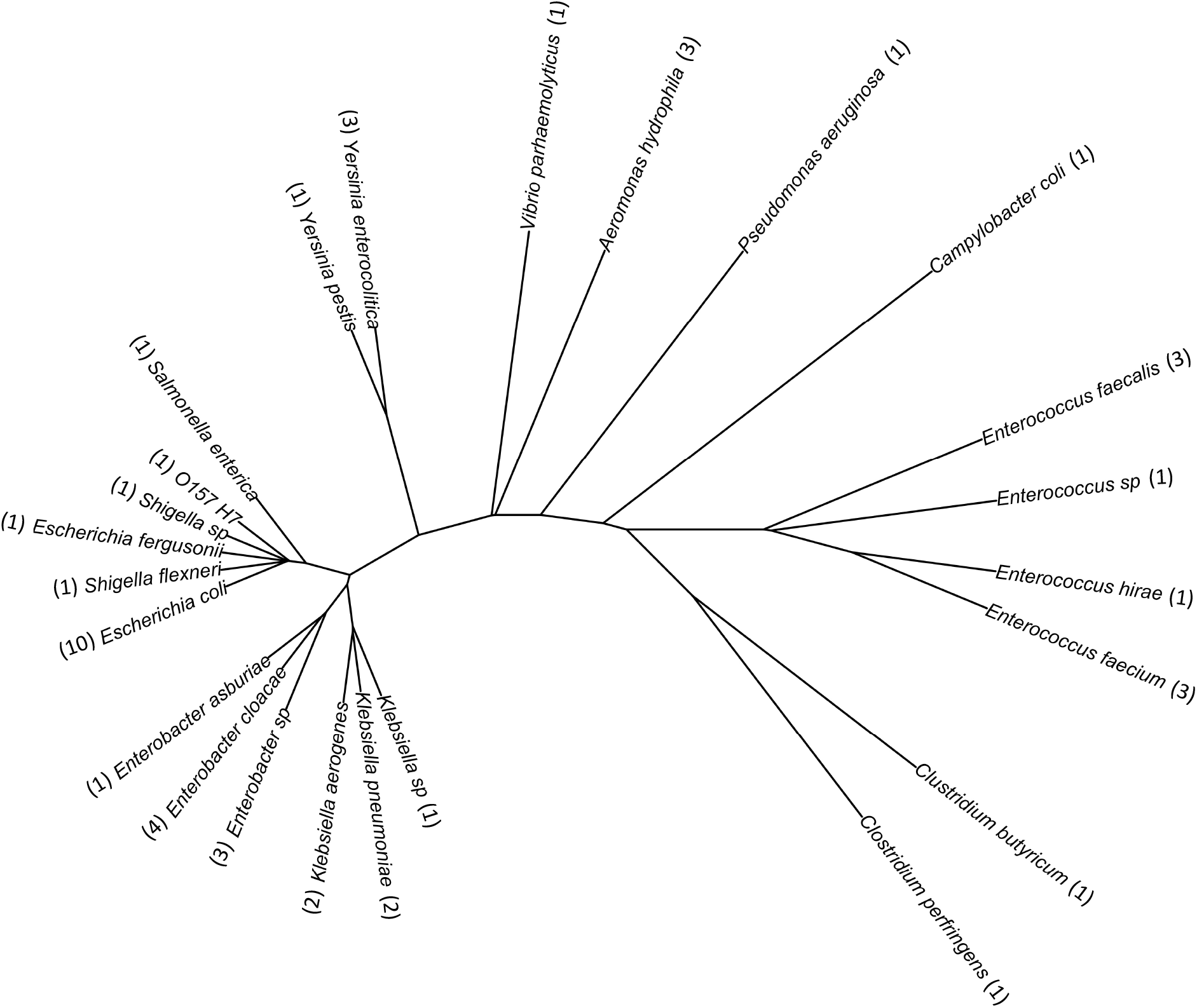
Taxonomic organization of enteric bacterial targets detected in studies included in the review. The number of manuscripts in which bacteria were detected are included in parenthesis where relevant.

The review revealed only three studies assessing enteric viruses in urban outdoor aerosols. Viral detections include human adenovirus and human adenovirus C; enterovirus A,B and C; and rotavirus A^58– 60^ Generally, the range of enteric microbes identified in bioaerosols in urban settings is small and we see an even smaller range represented among the culturable targets across studies. A complete breakdown by taxonomic classification of enteric microbes detected in the reviewed studies by all methods is in Table 2.

**Table 2:**
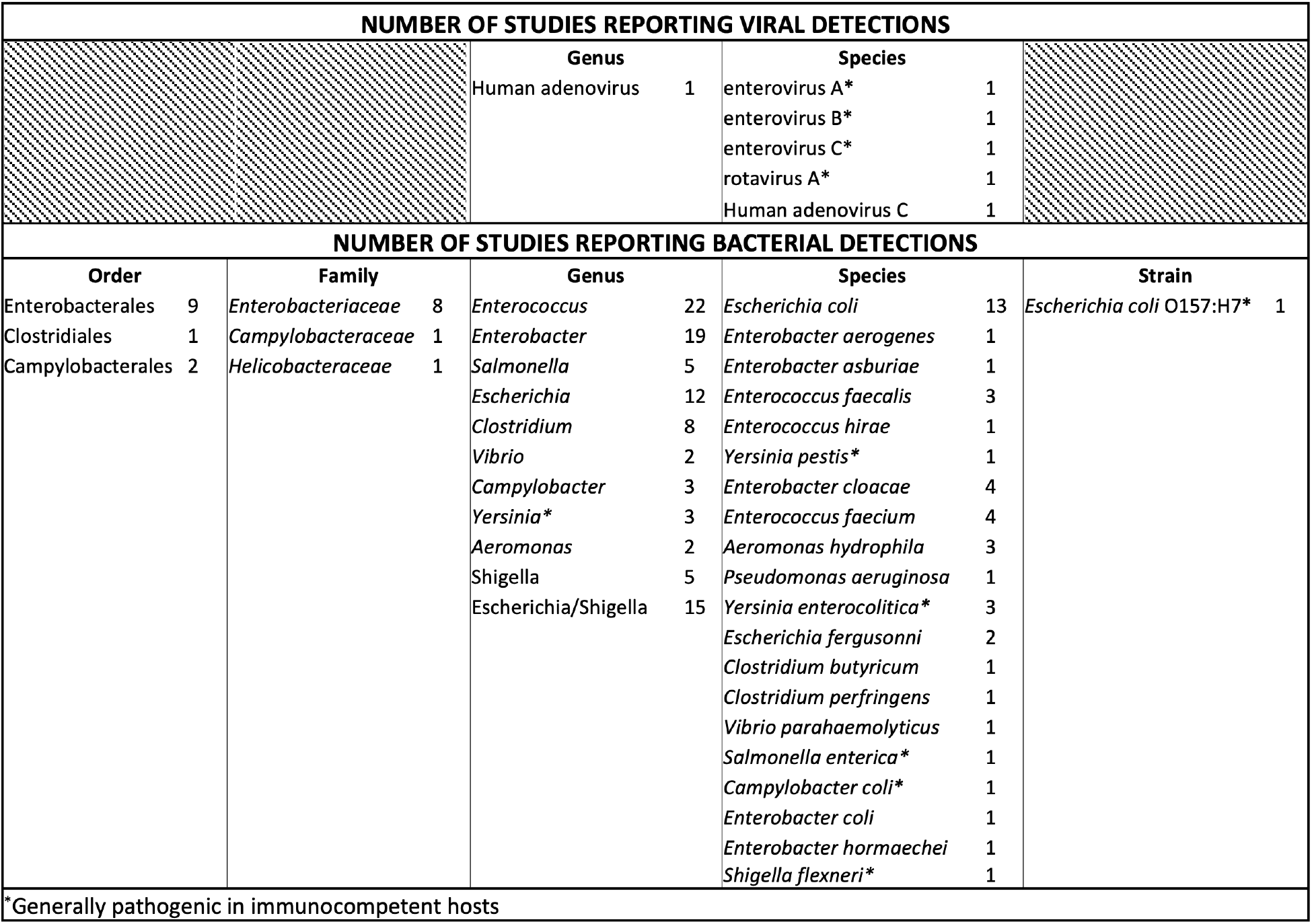
Number of studies reporting enteric microbe (commensal and pathogenic) detections broken down by taxonomic classification.

### 3.4 Antibiotic resistance

We identified fewer studies reporting on antibiotic resistant bacteria and antibiotic resistance encoding genes (ARGs) in bioaerosols (n=28). Using selective culture and often using disk diffusion methods,^120^ studies reported viable bacterial resistance to a broad range of antibiotic groups also highlighted on the US Food and Drug Administration’s, National Antimicrobial Resistance Monitoring System as existent or emerging threats,^121^ Some of these groups include β--lactams (penicillins, cephalosporins and carbapenems), tetracyclines, quinolones, aminoglycosides, sulfonamides, and phenicols. Through qPCR, a wide range of AR related genetic targets have been detected in urban aerosols such as antibiotic resistance genes (ARGs), mobile genetic elements (MGEs) encoding horizontal gene transfer ability, or mobile integrons (MIs) encoding the ability to undergo recombination and functional conversion of ARGs (Table 3).^122–124^ However, of the studies that detected ARGs in bioaerosols via PCR, 79% took place in upper-middle and high-income countries.

**Table 3:**
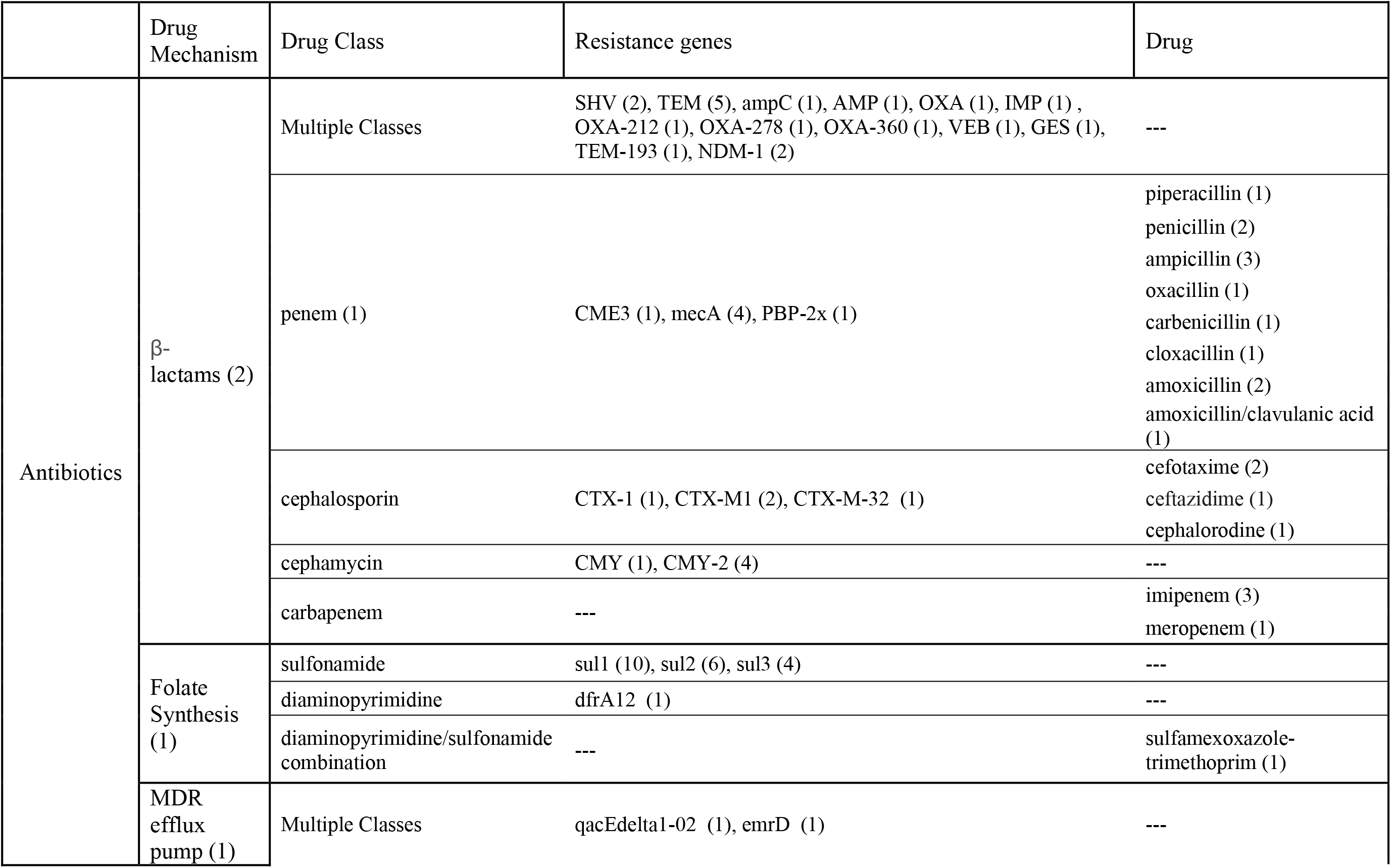
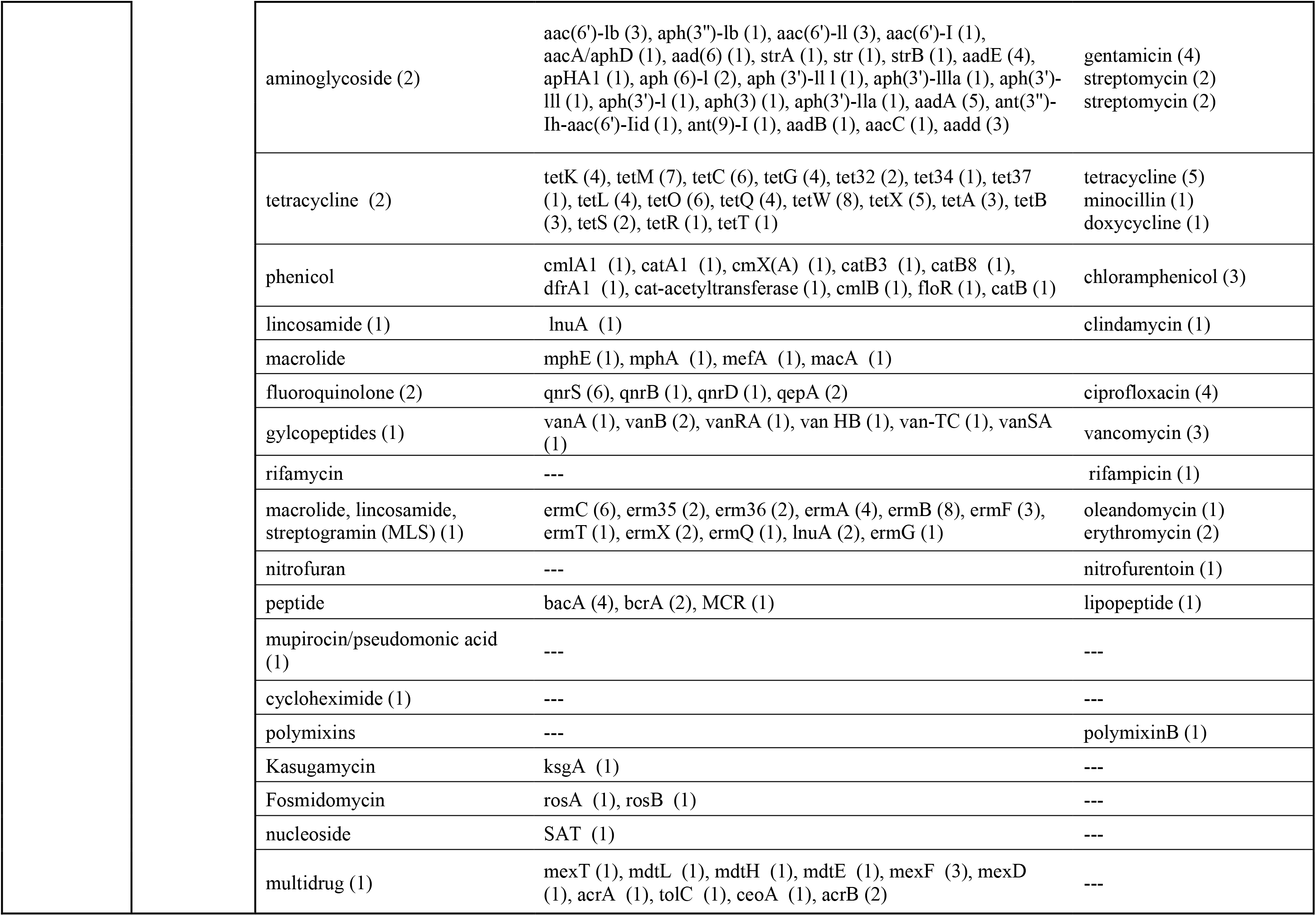
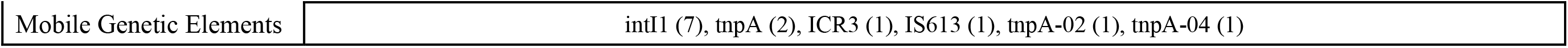
Antibiotic Resistance related categorization with number of studies citing each target (at various levels) in parenthesis.

|  | Drug Mechanism | Drug Class | Resistance genes | Drug |
| --- | --- | --- | --- | --- |
| Antibiotics | β-lactams (2) | Multiple Classes | SHV (2), TEM (5), ampC (1), AMP (1), OXA (1), IMP (1), OXA-212 (1), OXA-278 (1), OXA-360 (1), VEB (1), GES (1), TEM-193 (1), NDM-1 (2) | --- |
|  |  | penem (1) | CME3 (1), mecA (4), PBP-2x (1) | piperacillin (1)<br>penicillin (2)<br>ampicillin (3)<br>oxacillin (1)<br>carbenicillin (1)<br>cloxacillin (1)<br>amoxicillin (2)<br>amoxicillin/clavulanic acid (1) |
|  |  | cephalosporin | CTX-1 (1), CTX-M1 (2), CTX-M-32 (1) | cefotaxime (2)<br>ceftazidime (1)<br>cephalorodine (1) |
|  |  | cephamycin | CMY (1), CMY-2 (4) | --- |
|  |  | carbapenem | --- | imipenem (3)<br>meropenem (1) |
|  | Folate Synthesis (1) | sulfonamide | sul1 (10), sul2 (6), sul3 (4) | --- |
|  |  | diaminopyrimidine | dfrA12 (1) | --- |
|  |  | diaminopyrimidine/sulfonamide combination | --- | sulfamexoxazole-trimethoprim (1) |
|  | MDR efflux pump (1) | Multiple Classes | qacEdelta1-02 (1), emrD (1) | --- |

|  |  |  |  |
| --- | --- | --- | --- |
|  | aminoglycoside (2) | aac(6')-Ib (3), aph(3'')-Ib (1), aac(6')-II (3), aac(6')-I (1), aacA/aphD (1), aad(6) (1), strA (1), str (1), strB (1), aadE (4), apHA1 (1), aph (6)-I (2), aph (3')-II I (1), aph(3')-IIIa (1), aph(3')-III (1), aph(3')-I (1), aph(3) (1), aph(3')-IIa (1), aadA (5), ant(3'')-Ih-aac(6')-Iid (1), ant(9)-I (1), aadB (1), aacC (1), aadd (3) | gentamicin (4)<br>streptomycin (2)<br>streptomycin (2) |
|  | tetracycline (2) | tetK (4), tetM (7), tetC (6), tetG (4), tet32 (2), tet34 (1), tet37 (1), tetL (4), tetO (6), tetQ (4), tetW (8), tetX (5), tetA (3), tetB (3), tetS (2), tetR (1), tetT (1) | tetracycline (5)<br>minocycline (1)<br>doxycycline (1) |
|  | phenicol | cmlA1 (1), catA1 (1), cmX(A) (1), catB3 (1), catB8 (1), dfrA1 (1), cat-acetyltransferase (1), cmlB (1), floR (1), catB (1) | chloramphenicol (3) |
|  | lincosamide (1) | lnuA (1) | clindamycin (1) |
|  | macrolide | mphE (1), mphA (1), mefA (1), macA (1) |  |
|  | fluoroquinolone (2) | qnrS (6), qnrB (1), qnrD (1), qepA (2) | ciprofloxacin (4) |
|  | glycopeptides (1) | vanA (1), vanB (2), vanRA (1), van HB (1), van-TC (1), vanSA (1) | vancomycin (3) |
|  | rifamycin | --- | rifampicin (1) |
|  | macrolide, lincosamide, streptogramin (MLS) (1) | ermC (6), erm35 (2), erm36 (2), ermA (4), ermB (8), ermF (3), ermT (1), ermX (2), ermQ (1), lnuA (2), ermG (1) | oleandomycin (1)<br>erythromycin (2) |
|  | nitrofurantoin | --- | nitrofurantoin (1) |
|  | peptide | bacA (4), bcrA (2), MCR (1) | lipopeptide (1) |
|  | mupirocin/pseudomonic acid (1) | --- | --- |
|  | cycloheximide (1) | --- | --- |
|  | polymyxins | --- | polymyxinB (1) |
|  | Kasugamycin | ksgA (1) | --- |
|  | Fosmidomycin | rosA (1), rosB (1) | --- |
|  | nucleoside | SAT (1) | --- |
|  | multidrug (1) | mexT (1), mdtL (1), mdtH (1), mdtE (1), mexF (3), mexD (1), acrA (1), tolC (1), ceoA (1), acrB (2) | --- |

|  |  |
| --- | --- |
| Mobile Genetic Elements | intI1 (7), tnpA (2), ICR3 (1), IS613 (1), tnpA-02 (1), tnpA-04 (1) |

## 4.0 DISCUSSION

Results of this systematic review suggest that fecal signatures are present in ambient urban air across the globe, along with evidence suggesting aerosols may play a role in urban AR spread. Notably, studies conducted in cities in LMICs show a broad and diverse range of airborne microbial gene targets of possibly human fecal origin, potentially related to sanitation infrastructure deficits^125^ that permit uncontained waste flows in densely populated urban areas^44^. However, the results also reveal large gaps in the characterization and detection of important enteric microbes and AR in bioaerosols in ambient urban air. An especially concerning gap is that LMICs make up a minority of all study locations within the scope of this review, despite the fact that cities in these countries may be characterized by inadequate sanitation infrastructure and high population density, leading to widespread fecal contamination and disproportionately bearing the global burden of diarrheal disease, the cause of 1·8 million deaths worldwide in 2017.^126^ Given widespread carriage and shedding of important enteric pathogens, poor containment of sanitary waste near population centers, and the limited evidence that enteric pathogen-associated nucleic acids are widely present in aerosols, these sites should be an explicit focus of future monitoring efforts to understand the extent and implications of fecal microbe transport in aerosols.

Studies that do assess enteric microbes and AR are dominated by quantal (presence/absence) or semi-quantitative data obtained through sequencing. Such estimates target abundance in terms of the microbial community within each sample, as opposed to the environment in which the communities exist. While sequencing is useful in broadly characterizing and understanding infectious diseases and microbial communities in general, it is less useful in the context of transport phenomena and exposure measurements without additional contextual data, typically quantitative measurements that report density of specific genes or microbes per unit volume or mass.^127^ In the case of whole-genome sequencing, selection bias may be introduced if the microorganisms chosen for analysis are not representative of the broader population in the sample.^127^ When entire communities are sequenced, selection bias is resolved to a degree but additional shortcomings remain or are introduced such as quality control, quantification limitations, high limits of detection or lack of comparability across other methods and studies.^128^ Furthermore, characterizing specific genes such as those encoding antibiotic resistance requires increased sequencing depth which is not always possible with low concentration samples like bioaerosols.^129^ The studies that do quantitatively assess gene targets of enteric microbes (n=two) and AR (n=14) in bioaerosols provide more context, assessing targets in relation to the environmental media in which they are found as opposed to in relation to other targets present. Importantly, many of the most critical AR pathogens globally are known to be enriched in fecal wastes, including *E. coli, Klebsiella* spp., and *Pseudomonas aeruginosa*.^130^ All of these studies leverage real-time qPCR which is limited to interpretation based on standard curves as opposed to absolute quantification which allows for lower limits of detection^131^. While molecular detections are important in characterizing enteric microbes including pathogens of concern, detection of microbe-specific nucleic acid sequences do not indicate viability. Co-culture with fecal indicator bacteria most often detected in these studies (*E. coli, Enterobacter sp*., or *Enterococcus sp*.) may indirectly indicate potential viability of other microbes; vegetative bacteria are typically a conservative proxy for pathogen viability since they are readily inactivated in aerosols^132,133^. The combination of absolute quantification with an expansion of viability detection methods would contribute to advancing understanding of the potential public health relevance of this poorly understood pathway of transmission.^131^ The need for data characterizing aerosol transport of enteric pathogens and AR is most critical in settings where the risks of sanitation-related infections and AR spread are highest, typically LMICs where economic resources and laboratory capacity may be most limited. Although high-volume sample collection and methodologically sophisticated pathogen assays may often be unavailable in such settings, affordable and accessible detection of culturable fecal indicator bacteria and phenotypic resistance testing can provide valuable insight into potential health impacts for exposed populations.^58,134,135^

Studies included in this review reporting on enteric microbes including pathogens in bioaerosols in urban settings targeted a limited range of enteric microbes, with many studies (38%) reporting detection of commonly used fecal indicator bacteria (*Escherichia coli* and *Enteroccocus)*. Only one study in a tropical urban setting identified, through sequencing, the pathogenic strain of *Escherichia coli*, O157:H7 (EHEC) yielding a limited data pool that excludes other strains such as ETEC, EPEC and others that were responsible for over 60,000 deaths across the globe in 2016.^136^ Furthermore, though *Campylobacter coli* was detected in one study through sequencing at the species level, other pathogenic strains of *Campylobacter* remain uncharacterized in bioaerosols, even though it was responsible for 75,000 diarrheal deaths worldwide in 2016.^136^ Overall, the majority of enteric bacteria and viruses reported in the studies targeted and detected commensal enteric microbiota or opportunistic pathogens that typically only cause disease in immunocompromised individuals. Recent evidence reveals the growing relevance of opportunistic pathogens in clinical settings, particularly in the context of their low susceptibility to antibiotics and environmental origins^137^. Frank pathogens, which cause disease even in immunocompetent individuals are less represented in the review, making up less than half (39%) of all unique microbes reported at the strain and species levels across all studies (Table 2).

Few studies targeted and detected enteric viruses, even though fecal microbes are present and often are co-occurring with enteric viruses^44,138–140^. Viruses may be highly mobile in aerosols, but would not be captured in 16S rRNA sequencing that represents the bulk of analytical methods used in bioaerosols analysis. The available evidence from our review and other emerging evidence^44^ suggests that enteric viruses are important potential targets in urban aerosols, particularly norovirus given its well documented transport in aerosols^141–144^. Of the studies that employed methods that would be suited to the detection of enteric viruses (6 out of 101 covered in this review), 33% (n=2) identified them, including detections of human enterovirus, human rotavirus, and human adenovirus (Table 2).

No study included in our review targeted or reported enteric protozoans in bioaerosols in an urban setting, so an absence of evidence on detection of this class of enteric microbes should not be interpreted as evidence that they are not present in aerosols. These microbes tend to be larger in size (5-15 μm) and therefore experience rapid settling, and so may be of less interest in aerosol transport modeling, however. Despite this, some evidence suggests they could be widely detected in aerosols and may therefore merit further study. A study from rural Mexico detection of *Cryptosporidium* and *Giardia* in air samples via microscopy.^45^ A study published following our review reported detection of *Giardia duodenalis* and *Cryptosporidium* spp. near open sewers in both urban La Paz, Bolivia and Kanpur, India via qPCR, with

*G. duodenalis* detected in 22% of samples in La Paz and 18% of samples in Kanpur.^44^ In this study, *Cryptosporidium* spp. was detected in 9% and 3% of samples in La Paz and Kanpur, respectively, at average densities ranging from 9.3 to 560 gc/m^3^.^44^

The results of this review should be considered alongside a number of important limitations. First, though we used a double-blind search strategy and agreed *a priori* on inclusion criteria and other methods, bias or human error may have introduced some inconsistency in including literature for the review. Though we used a consensus-based approach to inclusion of studies, we may have missed or mischaracterized individual studies given the large number of papers we reviewed. Second, we excluded studies that were not translatable to English or that had non-extractable data that may have been valuable contributions within this scope of work. Third, because of the broad range of analysis methods, diagnostic tools, and variability in interpretation of data, we did not perform a structured meta-analysis and limited our results and discussion to a broad characterization and categorization of the literature. Future reviews of an expanded number of papers may be able to quantitatively synthesize and compare study findings according to methods and metadata using robust statistical methods well suited to meta-analysis.

## 5.0 CONCLUSION

Based on our interpretation of the results of this review, we can offer the following conclusions. First, enteric microbes and AR are widely detected in outdoor urban aerosols. A wide range of targets have been detected, though bacteria are most prominent; most analytical methods in the studies covered in this review employed methods that limited detection to bacteria only. Further attention on specific pathogens of interest and especially studies targeting enteric viruses and protozoa are needed. Second, we observe major gaps in evidence around microbial viability and quantitative estimation of specific enteric pathogens of global risk relevance. Quantitative data are needed alongside aerosol size data to advance fate and transport modeling, and viability of pathogens – even if indirect via bacterial indicator co-culture – is critically important to any assessment of the public health relevance of pathogen detection in aerosols. Third, we can conclude that important gaps in evidence are apparent specifically in settings where the risks of enteric pathogen exposure and AR emergence and spread are most acute: cities of LMICs, which are characterized in part by poor sanitation and high population density in close proximity to potential sources of bioaerosols containing enteric targets. We propose that future studies should address these gaps so that we will be able to better understand the potential for enteric microbes and AR in aerosols to impact global public health and to inform control strategies that can limit exposures when and where they are most critical.

## Data Availability

All data utilized in the present review are included in the supplemental material.

## 6.0 CONTRIBUTIONS

**Olivia Ginn**: Conceptualization, Methodology, Formal analysis, Literature Search, Data Curation and Verification, Writing- Original Draft, Writing- Review and Editing, Visualization, Supervision, Project Administration

**Sarah Lowry**: Methodology, Formal Analysis, Literature Search, Data Curation and Verification, Writing- Original Draft, Writing- Review and Editing

**Joe Brown**: Conceptualization, Methodology, Formal Analysis, Data Verification, Writing- Review and Editing, Funding Acquisition

