## Supplemental Material for "A systematic review of enteric pathogens and antibiotic resistance genes in outdoor urban aerosols"

**Supplementary Material to: A systematic review of enteric pathogens and antibiotic resistance genes in extramural urban aerosols**

Table S1. Prisma checklist.

| **Section/topic** | **#** | **Checklist item** | **Reported on page #** |
| --- | --- | --- | --- |
| **TITLE** | | |  |
| Title | 1 | Identify the report as a systematic review, meta-analysis, or both. | 10 |
| **ABSTRACT** | | |  |
| Structured summary | 2 | Provide a structured summary including, as applicable: background; objectives; data sources; study eligibility criteria, participants, and interventions; study appraisal and synthesis methods; results; limitations; conclusions and implications of key findings; systematic review registration number. | 10 |
| **INTRODUCTION** | | |  |
| Rationale | 3 | Describe the rationale for the review in the context of what is already known. | 12 |
| Objectives | 4 | Provide an explicit statement of questions being addressed with reference to participants, interventions, comparisons, outcomes, and study design (PICOS). | 14 |
| **METHODS** | | |  |
| Protocol and registration | 5 | Indicate if a review protocol exists, if and where it can be accessed (e.g., Web address), and, if available, provide registration information including registration number. | 14 |
| Eligibility criteria | 6 | Specify study characteristics (e.g., PICOS, length of follow-up) and report characteristics (e.g., years considered, language, publication status) used as criteria for eligibility, giving rationale. | 14 |
| Information sources | 7 | Describe all information sources (e.g., databases with dates of coverage, contact with study authors to identify additional studies) in the search and date last searched. | 14 |
| Search | 8 | Present full electronic search strategy for at least one database, including any limits used, such that it could be repeated. | Appendix A |
| Study selection | 9 | State the process for selecting studies (i.e., screening, eligibility, included in systematic review, and, if applicable, included in the meta-analysis). | 15 |
| Data collection process | 10 | Describe method of data extraction from reports (e.g., piloted forms, independently, in duplicate) and any processes for obtaining and confirming data from investigators. | 17 |
| Data items | 11 | List and define all variables for which data were sought (e.g., PICOS, funding sources) and any assumptions and simplifications made. | 17, Appendix A |
| Risk of bias in individual studies | 12 | Describe methods used for assessing risk of bias of individual studies (including specification of whether this was done at the study or outcome level), and how this information is to be used in any data synthesis. | n/a |
| Summary measures | 13 | State the principal summary measures (e.g., risk ratio, difference in means). | n/a |
| Synthesis of results | 14 | Describe the methods of handling data and combining results of studies, if done, including measures of consistency (e.g., I^2^) for each meta-analysis. | n/a |
| Risk of bias across studies | 15 | Specify any assessment of risk of bias that may affect the cumulative evidence (e.g., publication bias, selective reporting within studies). | n/a |
| Additional analyses | 16 | Describe methods of additional analyses (e.g., sensitivity or subgroup analyses, meta-regression), if done, indicating which were pre-specified. | 17 |
| **RESULTS** | | |  |
| Study selection | 17 | Give numbers of studies screened, assessed for eligibility, and included in the review, with reasons for exclusions at each stage, ideally with a flow diagram. | 17 |
| Study characteristics | 18 | For each study, present characteristics for which data were extracted (e.g., study size, PICOS, follow-up period) and provide the citations. | 18 |
| Risk of bias within studies | 19 | Present data on risk of bias of each study and, if available, any outcome level assessment (see item 12). | n/a |
| Results of individual studies | 20 | For all outcomes considered (benefits or harms), present, for each study: (a) simple summary data for each intervention group (b) effect estimates and confidence intervals, ideally with a forest plot. | n/a |
| Synthesis of results | 21 | Present results of each meta-analysis done, including confidence intervals and measures of consistency. | n/a |
| Risk of bias across studies | 22 | Present results of any assessment of risk of bias across studies (see Item 15). | n/a |
| Additional analysis | 23 | Give results of additional analyses, if done (e.g., sensitivity or subgroup analyses, meta-regression [see Item 16]). | 19 |
| **DISCUSSION** | | |  |
| Summary of evidence | 24 | Summarize the main findings including the strength of evidence for each main outcome; consider their relevance to key groups (e.g., healthcare providers, users, and policy makers). | 20 |
| Limitations | 25 | Discuss limitations at study and outcome level (e.g., risk of bias), and at review-level (e.g., incomplete retrieval of identified research, reporting bias). | 20 |
| Conclusions | 26 | Provide a general interpretation of the results in the context of other evidence, and implications for future research. | 22 |
| **FUNDING** | | |  |
| Funding | 27 | Describe sources of funding for the systematic review and other support (e.g., supply of data); role of funders for the systematic review. | 27 |

**Prisma flow diagram**

We include the original PRISMA flow diagram documenting the steps that lead to the final sample size included in the literature review.

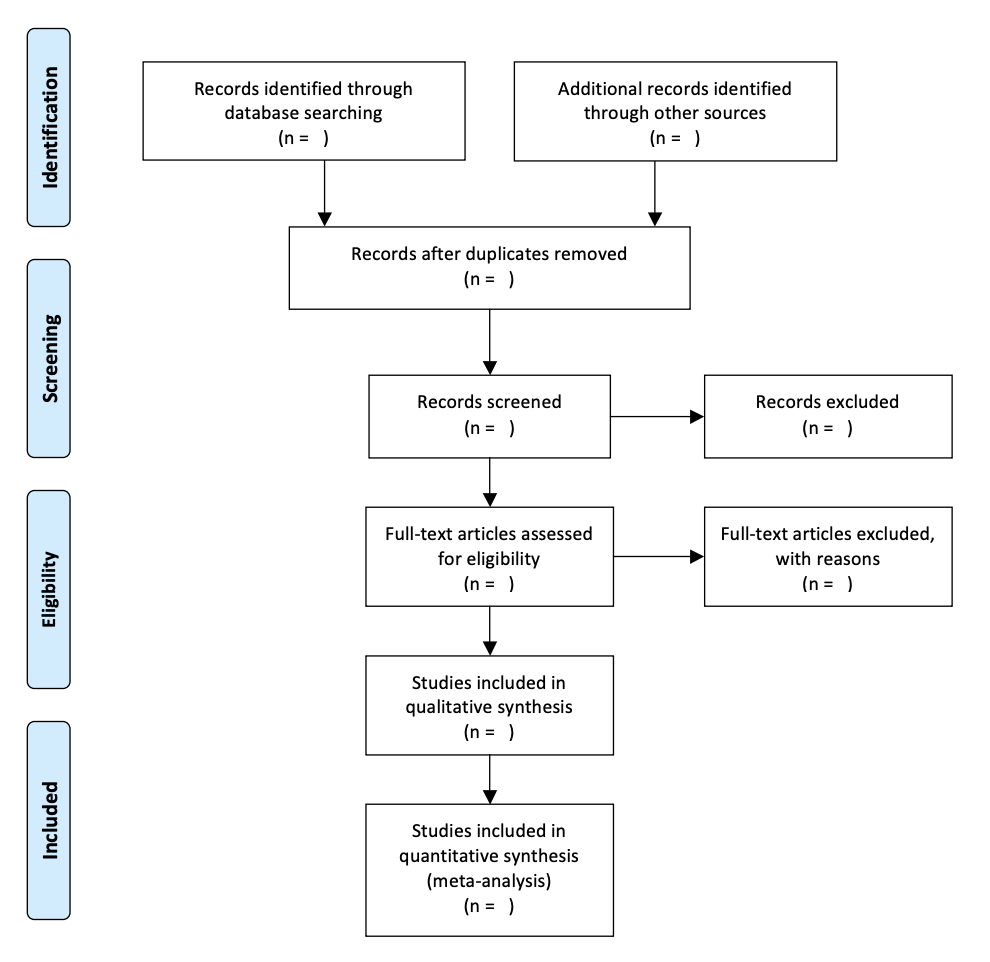

Figure S1. From Moher D, Liberati A, Tetzlaff J, Altman DG, The PRISMA Group (2009). *P*referred *R*eporting *I*tems for *S*ystematic Reviews and *M*eta-*A*nalyses: The PRISMA Statement. PLoS Med 6(7): e1000097. doi:10.1371/journal.pmed1000097

**Keyword search strategy**

To illustrate the complete search string strategy, we include the version used to search ISI Web of Science database below:

“TS=((bioaerosol OR  aerosol  OR  air  OR  "particulate  matter"  OR  PM  OR  airborne  or  aeromicrobiolog*  OR  aerosoliz*  OR  "bubbles  burst*"  OR  aeration  OR  "raindrop  impaction"  OR  "rain  impaction"  OR  re-aerosolized  OR  "air  sampling"  OR  "air  filter"  OR  "dry  filter"  OR  "dry  air  filter"  OR  impinger  OR  "passive  sampling"  OR  impaction  OR  "air  filtration"  OR  "active  air  sampling"  OR  "Anderson  Cascade  Impactor"  OR  impactor  OR  ACI  OR  SKC  OR  biosampler  OR  "sampling  cyclone"  OR  "Bi-Air  Filter  Cassette"  OR  "high-volume  air  sampling"  OR  "low-volume  air  sampling"  OR  "ACD-200  Bobcat  Air  Sampler"  OR  Bobcat  OR  "3100  Dry  Air  Sampler"  OR  "MD8  Air  Sampler"  OR  "Button  Air  Sampler"  OR  inhalation  OR  flux  OR  dispers*  OR  emission  OR  emit)  AND  (extramural OR outdoor* OR ambient OR outside OR atmosphere OR atmospheric)  AND  (urban OR cities OR city OR "dense population" OR "high population" OR "densely populated")  AND  ("enteric pathogen" OR enteric OR "intestinal pathogen" OR feces OR fecal OR "fecal indicator" OR coliform OR "fecal coliform" OR FIB OR FIO OR "enteric virus" OR adenovirus OR "adenovirus 4x" OR Aeromonas OR "Ascaris lumbricoides" OR astrovirus OR "Clostridium difficile" OR "C. difficile" OR Campylobacter OR "Campylobacter coli" OR "Campylobacter jejuni" OR Cryptosporidium OR "Cryptosporidium hominis" OR "Cryptosporidium parvum" OR "Entamoeba histolytica" OR "Escherichia coli" OR "E. coli" OR Shigella OR STEC OR "ST-ETEC" OR ETEC OR enterotoxigenic OR EIEC OR "O157:E7" OR EHEC OR EPEC OR EAEC OR Enterococcus OR "Enterococcus faecium" OR "Enterococcus faecalis" OR enterovirus* OR "Giardia lamblia" OR "Giardia spp." OR MS2 OR "MS2 phage" OR "Mycobacterium tuberculosis" OR "Norovirus GI" OR "Norovirus GII" OR "Rotavirus A" OR "Rotavirus B" OR "Rotavirus C" OR "Salmonella enterica" OR "Salmonella typhi" OR "typhoid" OR "Salmonella" OR "sapovirus" OR "Sapovirus I" OR "Sapovirus II" OR "Sapovirus IV" OR "Sapovirus V" OR "Shiga toxin" OR "Shiga-like toxin" OR "Trichuris trichiura" OR "Vibrio cholerae" OR "Yersinia spp." OR "Yersinia enterocolitica" OR STH OR "soil transmitted helminth" OR bacteriophage OR coliphage OR "Enterobius vermicularis" OR "Taenia spp." OR "Hymenolepis nana" OR "Strongyloides stercoralis" OR "Ancylostoma duodenale" OR "Necator americanus" OR "Helicobacter pylori" OR "H. pylori" OR "antimicrobial resistance" OR AMR OR ARG OR ARGs OR (antibiotic AND (resistance OR resistant))  OR  "antibiotic  resistant  genes"  OR  bcrR  OR  ermA  OR  ermB  OR  tetA  OR  tetC  OR  tetG  OR  tetM  OR  tetO  OR  tetQ  OR  tetW  OR  tetB  OR  tetL  OR  int1  OR  qnrS  OR  qnrB  OR  sul1  OR  sul2  OR  sul2  OR  aad  OR  vatE  OR  mexF  OR mphE OR  blaTEM  OR  ctx  OR  OXA-51  OR  OXA-23  OR  OXA-24  OR  OXA-58  OR  intl1  OR  blaOXA-51  OR  blaOXA-23  OR  blaOXA-24  OR  blaOXA-58  OR  aac  OR  metallo-beta-lactamase  OR  MBL  OR  oxacillinase  OR  MexAB-OprM  OR  MexCD-OprJ  OR  MexXY-OprM  OR  tetX  OR  blaSHA  OR  tnpA  OR  qepA  OR  blaCMY2  OR  erm35  OR  erm36  OR  ermC  OR  ermF  OR  ermT  OR  ermX  OR  tetK  OR  tet32  OR  tet37  OR  aac  OR  vanB  OR  vanRA  OR  vanSA  OR  aphA3  OR  mphA  OR  floR  OR  qacEdelta1  OR  dfrA1  OR  (((carbapenem OR tetracycline OR vancomycin OR sulfonamid OR quinolone OR macrolide OR multidrug OR aminoglycoside OR MLS OR β-lactam OR beta-lactam OR ESBL OR "extended spectrum β-lactam*" OR chloramphenicol OR bacitracin OR amikacin OR gentamicin OR kanamycin OR streptomycin OR "amoxicillin-clavulanic acid" OR "piperacillin-tazobactam" OR cefepime OR cefotaxime OR cefoxitin OR ceftazidime OR ceftiofur OR ceftriaxone OR cephalothin OR sulfamethoxazole OR sulfisoxazole OR trimethoprim-sulfamethoxazole OR azithromycin OR aztreonam OR imipenem OR meropenem OR ampicillin OR ciprofloxacin OR "nalidixic acid" OR cotrimoxazole OR cyclophosphamide OR sulbactam OR cefoperazone OR fluoroquinolone OR pandrug OR colistin OR polymyxin OR ofloxacin OR enrofloxacin)  NEAR  (resistance OR resistant) )  OR  MDR  OR  FCA  OR  MLSB)))

Table S2: Study data summary

| **Identify** | **Cat 1** | **Cat 2** | **Cat 3** | **First Author** | **Year Published** | **Location (Country)** | **Country GDP** | **Target** | **Sequencing Level** | **Detects** | **Reference** |
| --- | --- | --- | --- | --- | --- | --- | --- | --- | --- | --- | --- |
| forwards | AR | semi-quant | sequencing | Abd Aziz | 2018 | South Korea | high | All bacterial associated ARGs | --- | multidrug resistance efflux pumps, beta lactamase, resistance to fluoroquinolones | ^1^ |
| backwards | AR | quant | pcr | Echeverria-Palencia | 2017 | USA | high | Specific AR | --- | sul1, bla_shv | ^2^ |
| initial | AR | semi_quant | sequencing | Gandolfi | 2011 | Italy | high | Specific AR | --- | vanA, ermA, ermC, tetK, tetM, mecA | ^3^ |
| initial | AR | quant | culture | Gandolfi | 2011 | Italy | high | Specific AR | --- | enterococcus and actinobacter resistance to multiple abx | ^3^ |
| initial | AR | quant | pcr | Gao | 2016 | China | mid | Specific AR | --- | tetC, tetG, sul2, ermC | ^4^ |
| forwards | AR | semi_quant | pcr | Gat | 2017 | Israel | mid | Specific AR | --- | qnrS, sul1, intl1 | ^5^ |
| forwards | AR | semi_quant | sequencing | He | 2020 | China | mid | All bacterial associated ARGs | --- | heat map of ARG subtypes | ^6^ |
| forwards | AR | semi_quant | sequencing | Hu | 2018 | China | mid | All bacterial associated ARGs | --- | Many ARGs | ^7^ |
| backwards | AR | quant | culture | Kellogg | 2004 | Mali | low | Specific AR | --- | penicillin g, gentamicin, cefataxime, ciprofloxacin, erythromycin, chloramphenicol, tetracycline, ampicillin, nitrofurentoin, oxacillin | ^8^ |
| initial | AR | quant | pcr | Kumar | 2016 | India | low | Specific AR | --- | mecA | ^9^ |
| initial | AR | semi_quant | sequencing | Li | 2018 | Multiple | --- | All bacterial associated ARGs | --- | 39 ARG subtypes conferring resistance to 7 antibiotic types | ^10^ |
| forwards | AR | semi_quant | sequencing | Liang | 2020 | China | mid | All bacterial associated ARGs | --- | 34 ARGs and 10 MGEs | ^11^ |
| backwards | AR | quant | pcr | Mahdy | 1997 | Saudi Arabia | high | Specific AR | --- | Resistance to Microbes, Tetracycline, cephaloridine, carbenicillin, minocin, cloxacillin, amoxycillin | ^12^ |
| not sure | AR | semi-quant | sequencing | Mao | 2019 | China | mid | All bacterial associated ARGs | --- | penicillins, β-Lactam/β-Lactamase inhibitor combinations, cephems, lipopeptides, glycopeptides, penems, macrolides, lincosamides, Macrolides/Lincosamides combinations, pseudomonic acid, fluoroquinolones, folate pathway inhibitors, tetracyclines, phenicol, and aminoglycosides | ^13^ |
| initial | AR | quant | pcr | Mazar | 2016 | Israel | high | Specific AR | --- | intl1, ctx-m-32, sul1, qnrS, mecA | ^14^ |
| initial | AR | quant | pcr | Ouyang | 2020 | China | mid | Specific AR | --- | tetM, tetO, tetW, tetK, tetX, ermB, ermQ, mphE, aph3'IIIa, aph3'IIa | ^15^ |
| forwards | AR | quant | culture | Pathak | 2020 | India | low | Specific AR | --- | Vancomycin, Erythromycin, Chloramphenicol, Ampicillin | ^16^ |
| initial | AR | quant | culture | Raisi | 2012 | Crete | high | Specific AR | --- | Streptomycin, Cycloheximide | ^17^ |
| initial | AR | quant | culture | Salazar | 2020 | Bolivia | low | Specific AR | --- | Amoxicillin, ciproflaxin, gentamicin, meropenem, sulfamethoxazole-trimethoprim, tetracycline | ^18^ |
| forwards | AR | quant | culture | Sivri | 2016 | Turkey | mid | Specific AR | --- | ciprofloxacin, penicillin, doxycycline, imipenem, tetracycline, streptomycin, amoxicillin/clavulanic acid, vancomycin, gentamicin, clindamycin | ^19^ |
| forwards | AR | quant | pcr | Sun | 2020 | China | mid | Specific AR | --- | sul1, intI1, aadd, qnrs, blactx-M1, mexF, ermG, tetW, and ermB | ^20^ |
| initial | AR | quant | pcr | Wang | 2019 | China | mid | Specific AR | --- | 19 ARG subtypes: qnrS, blaCMY, blaTEM, blaAMP, blaOXA, ermC, ermB, ermA, sul3, sul2, sul1, tetW, tetQ, tetA, tetO, tetC, tetG, tetM | ^21^ |
| initial | AR | quant | pcr | Xie | 2018 | China | mid | Specific AR | --- | ermB, tetW, qnrS, intl1 | ^22^ |
| initial | AR | quant | pcr | Xie | 2019 | China | mid | Specific AR | --- | 6 ARGS (ermB, tetW, qnrS, lnuA, blatem, sul1) and 3 MGEs (intl1, tnpA-02, tnpA-04) | ^23^ |
| initial | AR | qual | pcr | Yadav | 2015 | India | low | Specific AR | --- | mecA | ^24^ |
| initial | AR | semi_quant | sequencing | Yang | 2018 | China | mid | All bacterial associated ARGs | --- | heat map of ARG subtypes | ^25^ |
| forwards | AR | quant | pcr | Zhang | 2019 | China | mid | Specific AR | --- | 39 ARG subtypes and 2 MGEs (intI1, tnpA) | ^26^ |
| forwards | AR | quant | culture | Zhao | 2020 | China | mid | Specific AR | --- | Carbapenem (ampicillin and imipenem) and Polymyxins (polymixin B) Resistance | ^27^ |
| forwards | AR | quant | pcr | Zhao | 2020 | China | mid | Specific AR | --- | 68 targets conferring resistance to: 9 antibiotics (i.e., aminoglycosides, multidrug, macrolidelincosamide- streptogramin B, beta-lactams, sulfonamides, tetracycline, vancomycin, chloramphenicol, and others) and 4 MGEs (2 integrase genes and 2 transposase genes | ^27^ |
| forwards | enteric | semi_quant | sequencing | Abd Aziz | 2018 | South Korea | high | All microbial communities | Genus | Escherichia | ^1^ |
| forwards | enteric | qual | culture | Agarwal | 2016 | India | low | Specific enteric pathogens | Genus | Escherichia | ^28^ |
| initial | enteric | semi_quant | sequencing | Amarloei | 2020 | Iran | mid | All microbial communities | Genus | Enterococcus | ^29^ |
| initial | enteric | semi_quant | sequencing | Bertolini | 2013 | Italy | high | All microbial communities | Order | Enterobacteriales | ^30^ |
| initial | enteric | semi_quant | sequencing | Bowers | 2011 | US | high | All microbial communities | Order | Enterobacteriales | ^31^ |
| initial | enteric | semi_quant | sequencing | Bowers | 2013 | US | high | All microbial communities | Family | Enterobacteriaceae | ^32^ |
| initial | enteric | semi_quant | sequencing | Brodie | 2006 | US | high | All microbial communities | Family | Campylobacteracea (Arcobacter), Helicobacteraceae | ^33^ |
| forwards | enteric | semi_quant | sequencing | Calderón-Ezquerro | 2020 | Mexico | mid | All microbial communities | Genus | Enterococcus | ^34^ |
| backwards | enteric | semi_quant | sequencing | Cao | 2014 | China | mid | All microbial communities | Species | Enterobacteriaceae, human adenovirus C | ^35^ |
| forwards | enteric | semi_quant | sequencing | Cha | 2016 | South Korea | high | All microbial communities | Genus | Escherichia-Shigella | ^36^ |
| backwards | enteric | quant | culture | Chen | 2012 | China | mid | Specific enteric pathogens | Species | Enterobacter spp | ^37^ |
| Forwards | enteric | semi_quant | sequencing | Cuthbertson | 2017 | NSF | high | All microbial communities | Genus | Enterococcus | ^38^ |
| forwards | enteric | semi_quant | sequencing | Du | 2018 | China | mid | All microbial communities | Genus | Escherichia | ^39^ |
| initial | enteric | semi_quant | sequencing | Du | 2018 | China | mid | All microbial communities | Genus | Clostridium, Enterobacter | ^40^ |
| forwards | enteric | semi_quant | sequencing | Fan | 2019 | China | mid | All microbial communities | Genus | Clostridium, Vibrio, Campylobacter, Enterococcus, Eschrichia-Shigella | ^41^ |
| Initial | enteric | semi_quant | sequencing | Fan | 2019 | China | mid | All microbial communities | Genus | Enterobacter | ^42^ |
| initial | enteric | semi_quant | sequencing | Fang | 2007 | China | mid | All microbial communities | Genus | Enterococcus, Escherichia, Yersinia | ^43^ |
| forwards | enteric | semi_quant | sequencing | Fang | 2018 | China | mid | All microbial communities | Order | Enterobacteriales | ^44^ |
| backwards | enteric | semi_quant | sequencing | Fierer | 2008 | US | high | All microbial communities | Family | Enterobacteriaceae | ^45^ |
| backwards | enteric | semi_quant | sequencing | Franzetti | 2011 | Italy | high | All microbial communities | Family | Enterobacteriaceae | ^46^ |
| forwards | enteric | semi_quant | sequencing | Fykse | 2015 | NSF | high | All microbial communities | Genus | Enterococcus | ^47^ |
| initial | enteric | semi_quant | sequencing | Gandolfi | 2011 | Italy | high | All microbial communities | Genus | Genus Enterococcus; Family Enterobacteriaceae | ^3^ |
| forwards | enteric | semi_quant | sequencing | Gandolfi | 2015 | Italy | high | All microbial communities | Order | Enterobacteriales | ^48^ |
| initial | enteric | semi_quant | sequencing | Gangamma | 2014 | India | low | All microbial communities | Species | Enterobacter aerogenes, Enterobacter asburie, Enterococcus faecalis, Enterococcus hirae | ^49^ |
| initial | enteric | semi_quant | pcr | Gao | 2016 | China | mid | Specific enteric pathogens | Species | Yersinia pestic, E. cloacae, E faecium | ^4^ |
| initial | enteric | semi_quant | sequencing | Gao | 2017 | China | mid | All microbial communities | Species | Enterococcus faecium, Escherichia coli, Enterobacter cloacae, Aeromonas hydrophila, Clostridium butricum, Vibrio parahaemolyticus, Clostridum perfringens | ^50^ |
| backwards | enteric | semi_quant | sequencing | Garcia-Mena | 2016 | Mexico | mid | All microbial communities | Genus | Enterococcus, Escherichia, Enterobacteriaceae, Enterobacter | ^51^ |
| forwards | enteric | semi_quant | sequencing | Gat | 2017 | Israel | high | All microbial communities | Genus | Escherichia/Shigella | ^5^ |
| initial | enteric | semi_quant | sequencing | Gonzalez Martin | 2018 | Spain | high | Specific enteric pathogens | Species | Human Adenoviruses, Enteroviruses, and Rotaviruses | ^52^ |
| forwards | enteric | semi_quant | sequencing | Gou | 2016 | China | mid | All microbial communities | Genus | Enterobacter, Clostridium XI, Escherichia-Shigella, Clostridium sensu stricto, Klebsiella | ^53^ |
| forwards | enteric | semi_quant | sequencing | Hai | 2019 | Vietnam | low | All microbial communities | Genus | Enterobacteriacaea, Aeromonas | ^54^ |
| forwards | enteric | semi_quant | sequencing | Hu | 2020 | China | mid | All microbial communities | Species | Escherichia, Salmonella enterica | ^55^ |
| forwards | enteric | quant | culture | Huertas | 2018 | Columbia | mid | Specific enteric pathogens | Species | Enterococcus spp | ^56^ |
| initial | enteric | quant | culture | Hurtado | 2014 | Mexico | mid | Specific enteric pathogens | Genus | Enterococcus, Escherichia, Enterobacter | ^57^ |
| initial | enteric | semi_quant | sequencing | Innocente | 2017 | Italy | high | All microbial communities | Genus | Escherichia-Shigella | ^58^ |
| forwards | enteric | semi_quant | sequencing | Jang | 2018 | South Korea | high | All microbial communities | Order | Enterobacteriales, Clostridiales, Enterobacteriacea, Campylobacterales, Arcobacter | ^59^ |
| forwards | enteric | semi_quant | sequencing | Ji | 2019 | China | mid | All microbial communities | Species | Escherichia-Shigella, Enterococcus, Campylobacter coli | ^60^ |
| forwards | enteric | semi_quant | sequencing | Karlsson | 2020 | Sweden | high | All microbial communities | Order | Enterobacteriales | ^61^ |
| forwards | enteric | quant | culture | Kaushik | 2012 | Singapore | high | Specific enteric pathogens | Species | E. coli, P. aeruginosa, K. pneumonia and A. hydrophila | ^62^ |
| initial | enteric | semi_quant | sequencing | Li | 2018 | Multiple |  | All microbial communities | Genus | Escherichia/Shigella, Enterococcus | ^63^ |
| initial | enteric | semi_quant | sequencing | Li | 2019 | China | mid | All microbial communities | Species | Yersinia enterocolitica, enterococcus faecium | ^64^ |
| forwards | enteric | semi_quant | sequencing | Li | 2019 | China | mid | All microbial communities | Genus | Escherichia-Shigella | ^65^ |
| forwards | enteric | semi_quant | sequencing | Liang | 2020 | China | mid | All microbial communities | Genus | Escherichia-Shigella, Enterococcus, Salmonella | ^11^ |
| initial | enteric | semi_quant | sequencing | Lu | 2018 | China | mid | All microbial communities | Genus | Clostridium | ^66^ |
| forwards | enteric | semi_quant | sequencing | Mhuireach | 2019 | US | high | All microbial communities | Family | Enterobacteriaceae | ^67^ |
| initial | enteric | semi_quant | sequencing | Mu | 2020 | China | mid | All microbial communities | Genus | Escherichia-shigella, Enterococcus | ^68^ |
| initial | enteric | semi_quant | sequencing | Najafi | 2014 | Iran | mid | All microbial communities | Species | Escherichia coli, Enterobacter, Klebsiella | ^69^ |
| forwards | enteric | semi_quant | sequencing | Nanclares Castaneda | 2020 | Columbia | mid | All microbial communities | Genus | Enterobacter | ^70^ |
| initial | enteric | semi_quant | sequencing | Núñez | 2019 | Spain | mid | All microbial communities | Genus | Enterobacteriales, Aeromonas, Campylobacter, Enterobacter, Enterococcus, Escherichia-Shigella, Vibrio | ^71^ |
| forwards | enteric | semi_quant | sequencing | Núñez | 2020 | Spain | mid | All microbial communities | Genus | Enterobacter | ^72^ |
| initial | enteric | semi_quant | sequencing | Ouyang | 2020 | China | mid | All microbial communities | Order | Enterobacteriales | ^15^ |
| forwards | enteric | semi_quant | sequencing | Pan | 2019 | China | mid | All microbial communities | Order | Enterobacteriales | ^73^ |
| initial | enteric | quant | culture | Raghav | 2020 | India | low | Specific enteric pathogens | Species | Escherichia coli, Enterobacter aerogenes | ^74^ |
| initial | enteric | quant | culture | Rocha-Melogno | 2020 | Bolivia | low | Specific enteric pathogens | Species | E.coli, TC | ^75^ |
| initial | enteric | quant | pcr | Rocha-Melogno | 2020 | Bolivia | low | Specific enteric pathogens | Species | Human adenovirus and Human enterovirus | ^75^ |
| forwards | enteric | semi_quant | sequencing | Rodriguez-Gomez | 2020 | NSF | high | All microbial communities | Species | Enterobacter cloacae, Klebsiella sp. | ^76^ |
| forwards | enteric | semi_quant | sequencing | Romano | 2020 | Italy | high | All microbial communities | Genus | Enterobacter | ^77^ |
| initial | enteric | quant | culture | Rosas | 1997 | Mexico | mid | Specific enteric pathogens | Species | Escherichia coli | ^78^ |
| forwards | enteric | quant | pcr | Santos | 2019 | Mexico | mid | Specific enteric pathogens | Species | Enterococcus faecalis | ^79^ |
| initial | enteric | quant | culture | Santos-Burgoa | 1994 | Mexico | mid | Specific enteric pathogens | Species | E. coli, Enterobacter, S. faecalis (enterococci) | ^80^ |
| backwards | enteric | semi_quant | sequencing | Serrano-Silva | 2017 | Mexico | mid | All microbial communities | Family | Enterobacteriaceae | ^81^ |
| initial | enteric | semi_quant | sequencing | Shenn | 2019 | China | mid | All microbial communities | Genus | Enterococcus | ^82^ |
| initial | enteric | semi_quant | sequencing | Soleimani | 2016 | Iran | mid | All microbial communities | Genus | Enterococcus | ^83^ |
| backwards | enteric | semi_quant | sequencing | Soto | 2009 | Spain | mid | All microbial communities | Genus | Yersinia, Enterobacteria | ^84^ |
| forwards | enteric | semi_quant | sequencing | Stewart | 2020 | US | high | All microbial communities | Genus | Escherichia, Salmonella | ^85^ |
| forwards | enteric | semi_quant | sequencing | Sun | 2018 | China | mid | All microbial communities | Genus | Escherichia-Shigella, Enterococcus | ^86^ |
| forwards | enteric | semi_quant | sequencing | Sun | 2020 | China | mid | All microbial communities | Species | Escherichia-Shigella, Enterobacter, Yersinia enterocolitica, Shigella spp., Klebsiella pneumonia, Gardnerella vaginalis, Prevotella spp., Enterobacter spp. | ^20^ |
| initial | enteric | semi_quant | sequencing | Velez-Quinones | 2013 | US and Mali |  | All microbial communities | Species | Enterobacter spp, Aeromonas hydrophila | ^87^ |
| Backwards | enteric | semi_quant | sequencing | Wei | 2016 | China | mid | All microbial communities | Genus | Enterobacter, Enterococcus, Clostridium | ^88^ |
| forwards | enteric | semi_quant | sequencing | Wei | 2020 | China | mid | All microbial communities | Genus | Enterobacteriaceae, Aeromonas | ^89^ |
| forwards | enteric | semi_quant | sequencing | Woo | 2013 | China | mid | All microbial communities | Strain | Escherichia coli O157:H7, Shigella and Salmonella | ^90^ |
| initial | enteric | quant | culture | Wu | 2017 | China | mid | Specific enteric pathogens | Species | Yersinia enterocolitica, Salmonella, E. coli | ^91^ |
| forwards | enteric | semi_quant | sequencing | Xu | 2017 | China | mid | All microbial communities | Species | Clostridium, Escherichia, Escherichia fergusonii | ^92^ |
| initial | enteric | semi_quant | sequencing | Xu | 2017 | China | mid | All microbial communities | Genus | Escherichia-Shigella, Enterococcus, Enterobacter | ^93^ |
| initial | enteric | qual | culture | Yadav | 2015 | India | low | Specific enteric pathogens | Species | Enterococcus species, Escherichia coli | ^24^ |
| forwards | enteric | semi_quant | sequencing | Yan | 2018 | China | mid | All microbial communities | Genus | Enterobacter, clostridium | ^94^ |
| initial | enteric | semi_quant | sequencing | Yang | 2018 | China | mid | All microbial communities | Genus | Enterococcus, Escherichia | ^25^ |
| backwards | enteric | quant | culture | Yassin | 2010 | Kuwait | high | Specific enteric pathogens | Species | Escherichia coli | ^95^ |
| forwards | enteric | semi_quant | sequencing | Yooseph | 2013 | US | high | All microbial communities | Genus | Escherichia | ^96^ |
| backwards | enteric | semi_quant | sequencing | Zhang | 2019 | China | mid | All microbial communities | Genus | Shigella | ^97^ |
| forwards | enteric | semi_quant | sequencing | Zhao | 2020 | China | mid | All microbial communities | Genus | Escherichia-Shigella | ^27^ |
| forwards | enteric | semi_quant | sequencing | Zhen | 2017 | China | mid | All microbial communities | Order | Enterobacteriales | ^98^ |
| backwards | enteric | semi_quant | sequencing | Zhou | 2018 | China | mid | All microbial communities | Species | Enterobacter hormaechei, Enterobacter cloacae, Shigella flexneri | ^99^ |

1	Abd Aziz A, Lee K, Park B, *et al.* Comparative study of the airborne microbial communities and their functional composition in fine particulate matter (PM2.5) under non-extreme and extreme PM2.5 conditions. *Atmos Environ* 2018; **194**: 82–92.

2	Echeverria-Palencia CM, Thulsiraj V, Tran N, *et al.* Disparate Antibiotic Resistance Gene Quantities Revealed across 4 Major Cities in California: A Survey in Drinking Water, Air, and Soil at 24 Public Parks. *ACS Omega* 2017; **2**: 2255–63.

3	Gandolfi I, Franzetti A, Bertolini V, Gaspari E, Bestetti G. Antibiotic resistance in bacteria associated with coarse atmospheric particulate matter in an urban area. *J Appl Microbiol* 2011; **110**: 1612–20.

4	Gao X, Shao M, Luo Y, *et al.* Airborne bacterial contaminations in typical Chinese wet market with live poultry trade. *Sci Total Environ* 2016; **572**: 681–7.

5	Gat D, Mazar Y, Cytryn E, Rudich Y. Origin-Dependent Variations in the Atmospheric Microbiome Community in Eastern Mediterranean Dust Storms. *Environ Sci Technol* 2017; **51**: 6709–18.

6	He P, Wu Y, Huang W, *et al.* Characteristics of and variation in airborne ARGs among urban hospitals and adjacent urban and suburban communities: A metagenomic approach. *Environ Int* 2020; **139**. DOI:10.1016/j.envint.2020.105625.

7	Hu J, Zhao F, Zhang XX, *et al.* Metagenomic profiling of ARGs in airborne particulate matters during a severe smog event. *Sci Total Environ* 2018; **615**: 1332–40.

8	Kellogg CA, Griffin DW. Aerobiology and the global transport of desert dust. *Trends Ecol Evol* 2006; **21**: 638–44.

9	Kumar P, Goel AK. Prevalence of Methicillin Resistant Staphylococcal Bioaerosols in andaround Residential Houses in an Urban Area in Central India. *J Pathog* 2016; **2016**. DOI:10.1155/2016/7163615.

10	Li J, Cao J, Zhu YG, *et al.* Global Survey of Antibiotic Resistance Genes in Air. *Environ Sci Technol* 2018; **52**: 10975–84.

11	Liang Z, Yu Y, Ye Z, Li G, Wang W, An T. Pollution profiles of antibiotic resistance genes associated with airborne opportunistic pathogens from typical area, Pearl River Estuary and their exposure risk to human. *Environ Int* 2020; **143**: 105934.

12	Mahdy HM, El-Sehrawi MH. Airborne bacteria in the atmosphere of El-Taif region, Saudi Arabia. *Water Air Soil Pollut* 1997; **98**: 317–24.

13	Mao Y, Ding P, Wang Y, *et al.* Comparison of culturable antibiotic-resistant bacteria in polluted and non-polluted air in Beijing, China. *Environ Int* 2019; **131**: 104936.

14	Mazar Y, Cytryn E, Ere Y, Rudich Y. Effect of Dust Storms on the Atmospheric Microbiome in the EasternMediterranean. *Environ Sci Technol* 2016; **50**: 4194–202.

15	Ouyang W, Gao B, Cheng H, *et al.* Airborne bacterial communities and antibiotic resistance gene dynamicsin PM2.5 during rainfall. *Environ Int* 2020; **134**. DOI:10.1016/j.envint.2019.105318.

16	Pathak B, Borah D, Khataniar A, Bhuyan PK, Buragohain AK. Characterization of bioaerosols in Northeast India in terms of culturable biological entities along with inhalable, thoracic and alveolar particles. *J Earth Syst Sci* 2020; **129**. DOI:10.1007/s12040-020-01406-z.

17	Raisi L, Katsivela E, Lazaridis M, Aleksandropoulou V. Size distribution of viable, cultivable, airborne microbes and their relationship to particulate matter concentrations and meteorological conditions in a Mediterranean site [electronic resource]. *Aerobiologia (Bologna)* 2013; **29**: 233–48.

18	Salazar D, Ginn O, Brown J, Soria F, Garvizu C. Assessment of antibiotic resistant coliforms from bioaerosol samples collected above a sewage-polluted river in La Paz, Bolivia. *Int J Hyg Environ Health* 2020; **228**: 113494.

19	Sivri N, Bagcigil AF, Metiner K, *et al.* Culturable airborne bacteria and isolation of methicillin-resistantcoagulase-negative staphylococci from outdoor environments on Europeanside of Istanbul, Turkey. *Arch Environ Prot* 2016; **42**: 77–86.

20	Sun X, Li D, Li B, *et al.* Exploring the disparity of inhalable bacterial communities and antibiotic resistance genes between hazy days and non-hazy days in a cold megacity in Northeast China. *J Hazard Mater* 2020; **398**. DOI:10.1016/j.jhazmat.2020.122984.

21	Wang Y, Wang C, Song L. Distribution of antibiotic resistance genes and bacteria from sixatmospheric environments: Exposure risk to human. *Sci Total Environ* 2019; **694**. DOI:10.1016/j.scitotenv.2019.133750.

22	Xie J, Jin L, Luo X, Zhao Z, Li X. Seasonal Disparities in Airborne Bacteria and Associated Antibiotic Resistance Genes in PM2.5 between Urban and Rural Sites. *Environ Sci Technol Lett* 2018; **5**: 74–9.

23	Xie J, Jin L, He T, *et al.* Bacteria and Antibiotic Resistance Genes (ARGs) in PM2.5 from China: Implications for Human Exposure. *Environ Sci Technol* 2019; **53**: 963–72.

24	Yadav J, Kumar A, Mahor P, *et al.* Distribution of airborne microbes and antibiotic susceptibility patternof bacteria during Gwalior trade fair, Central India. *J Formos Med Assoc* 2015; **114**: 639–46.

25	Yang Y, Zhou R, Chen B, Zhang T, Hu L, Zou S. Characterization of airborne antibiotic resistance genes from typical bioaerosol emission sources in the urban environment using metagenomic approach. *Chemosphere* 2018; **213**: 463–71.

26	Zhang T, Li X, Wang M, *et al.* Time-resolved spread of antibiotic resistance genes in highly polluted air. *Environ Int* 2019; **127**: 333–9.

27	Zhao Y, Chen Z, Hou J, *et al.* Monitoring antibiotic resistomes and bacterial microbiomes in the aerosols from fine, hazy, and dusty weather in Tianjin, China using a developed high-volume tandem liquid impinging sampler. *Sci Total Environ* 2020; **731**: 139242.

28	Agarwal S, Mandal P, Srivastava A. Quantification and Characterization of Size-segregated Bioaerosols at Municipal Solid Waste Dumping Site in Delhi. *Procedia Environ Sci* 2016; **35**: 400–7.

29	Amarloei A, Fazlzadeh M, Jafari AJ, Zarei A, Mazloomi S. Particulate matters and bioaerosols during Middle East dust storms events in Ilam, Iran. *Microchem J* 2020; **152**. DOI:10.1016/j.microc.2019.104280.

30	Bertolini V, Innocente E, Rampazzo G, *et al.* Temporal variability and effect of environmental variables on airborne bacterial communities in an urban area of Northern Italy [electronic resource]. *Appl Microbiol Biotechnol* 2013; **97**: 6561–70.

31	Bowers RM, Sullivan AP, Costello EK, Collett JL, Knight R, Fierer N. Sources of bacteria in outdoor air across cities in the midwestern United States. *Appl Environ Microbiol* 2011; **77**: 6350–6.

32	Bowers RM, Clements N, Emerson JB, Wiedinmyer C, Hannigan MP, Fierer N. Seasonal Variability in Bacterial and Fungal Diversity of the Near-Surface Atmosphere. *Environ Sci Technol* 2013; **47**: 12097–106.

33	Brodie EL, DeSantis TZ, Parker JPM, Zubietta IX, Piceno YM, Andersen GL. Urban aerosols harbor diverse and dynamic bacterial populations. *Proc Natl Acad Sci* 2007; **104**: 299–304.

34	Calderón-Ezquerro MC, Serrano-Silva N, Brunner-Mendoza C. Metagenomic characterisation of bioaerosols during the dry season in Mexico City. *Aerobiologia (Bologna)* 2020; **36**: 493–505.

35	Cao C, Jiang W, Wang B, *et al.* Inhalable microorganisms in Beijing’s PM2.5 and PM10 pollutants during a severe smog event. *Environ Sci Technol* 2014; **48**: 1499–507.

36	Cha S, Lee D, Jang JH, Lim S, Yang D, Seo T. Alterations in the airborne bacterial community during Asian dust events occurring between February and March 2015 in South Korea. *Sci Rep* 2016; **6**: 1–9.

37	Chen X, Ran P, Ho K, *et al.* Concentrations and size distributions of airborne microorganisms in guangzhou during summer. *Aerosol Air Qual Res* 2012; **12**: 1336–44.

38	Cuthbertson L, Amores-Arrocha H, Malard LA, Els N, Sattler B, Pearce DA. Characterisation of arctic bacterial communities in the air above svalbard. *Biology (Basel)* 2017; **6**. DOI:10.3390/biology6020029.

39	Du P, Du R, Lu Z, Ren W, Fu P. Variation of bacterial and fungal community structures in PM2.5collected during the 2014 APEC summit periods. *Aerosol Air Qual Res* 2018; **18**: 444–55.

40	Du P, Du R, Ren W, Lu Z, Fu P. Seasonal variation characteristic of inhalable microbial communities in PM2.5 in Beijing city, China. *Sci Total Environ* 2018; **610**–**611**: 308–15.

41	Fan XY, Gao JF, Pan KL, Li DC, Dai HH, Li X. More obvious air pollution impacts on variations in bacteria than fungi and their co-occurrences with ammonia-oxidizing microorganisms in PM2.5. *Environ Pollut* 2019; **251**: 668–80.

42	Fan C, Li Y, Liu P, *et al.* Characteristics of airborne opportunistic pathogenic bacteria during autumn and winter in Xi’an, China. *Sci Total Environ* 2019; **672**: 834–45.

43	Fang Z, Ouyang Z, Zheng H, Wang X, Hu L. Culturable airborne bacteria in outdoor environments in Beijing, China. *Microb Ecol* 2007; **54**: 487–96.

44	Fang Z, Guo W, Zhang J, Lou X. Influence of heat events on the composition of airborne bacterial communities in urban ecosystems. *Int J Environ Res Public Health* 2018; **15**. DOI:10.3390/ijerph15102295.

45	Fierer N, Liu Z, Rodríguez-Hernández M, Knight R, Henn M, Hernandez MT. Short-term temporal variability in airborne bacterial and fungal populations. *Appl Environ Microbiol* 2008; **74**: 200–7.

46	Franzetti A, Gandolfi I, Gaspari E, Ambrosini R, Bestetti G. Seasonal variability of bacteria in fine and coarse urban air particulate matter. *Appl Microbiol Biotechnol* 2011; **90**: 745–53.

47	Fykse EM, Tjärnhage T, Humppi T, *et al.* Identification of airborne bacteria by 16S rDNA sequencing, MALDI-TOF MS and the MIDI microbial identification system. *Aerobiologia (Bologna)* 2015; **31**: 271–81.

48	Gandolfi I, Bertolini V, Bestetti G, *et al.* Spatio-temporal variability of airborne bacterial communities and their correlation with particulate matter chemical composition across two urban areas. *Appl Microbiol Biotechnol* 2015; **99**: 4867–77.

49	Gangamma S. Characteristics of airborne bacteria in Mumbai urban environment. *Sci Total Environ* 2014; **488**–**489**: 70–4.

50	Gao J-F, Fan X-Y, Li H-Y, Pan K-L. Airborne Bacterial Communities of PM2.5 in Beijing-Tianjin-HebeiMegalopolis, China as Revealed By Illumina MiSeq Sequencing: A CaseStudy. *AEROSOL AIR Qual Res* 2017; **17**: 788–98.

51	García-Mena J, Murugesan S, Pérez-Muñoz AA, *et al.* Airborne Bacterial Diversity from the Low Atmosphere of Greater Mexico City. *Microb Ecol* 2016; **72**: 70–84.

52	Gonzalez-Martin C, Coronado-Alvarez NM, Teigell-Perez N, *et al.* Analysis of the Impact of African Dust Storms on the Presence of EntericViruses in the Atmosphere in Tenerife, Spain. *AEROSOL AIR Qual Res* 2018; **18**: 1863–73.

53	Gou H, Lu J, Li S, Tong Y, Xie C, Zheng X. Assessment of microbial communities in PM1 and PM10 of Urumqi during winter. *Environ Pollut* 2016; **214**: 202–10.

54	Hai VD, Hoang SMT, Hung NTQ, *et al.* Characteristics of airborne bacteria and fungi in the atmosphere in Ho Chi Minh city, Vietnam - A case study over three years. *Int Biodeterior Biodegrad* 2019; **145**: 104819.

55	Hu Z, Liu H, Zhang H, *et al.* Temporal discrepancy of airborne total bacteria and pathogenic bacteria between day and night. *Environ Res* 2020; **186**: 109540.

56	Huertas ME, Acevedo-Barrios RL, Rodríguez M, Gaviria J, Arana R, Arciniegas C. Identification and Quantification of Bioaerosols in a Tropical Coastal Region: Cartagena de Indias, Colombia. *Aerosol Sci Eng* 2018; **2**: 206–15.

57	Hurtado L, Rodriguez G, Lopez J, *et al.* Characterization of atmospheric bioaerosols at 9 sites in Tijuana, Mexico. *Atmos Environ* 2014; **96**: 430–6.

58	Innocente E, Franzetti A, Facca C, *et al.* Influence of seasonality, air mass origin and particulate matter chemical composition on airborne bacterial community structure in the Po Valley, Italy. *Sci Total Environ* 2017; **593**–**594**: 677–87.

59	Jang G Il, Hwang CY, Cho BC. Effects of heavy rainfall on the composition of airborne bacterial communities. *Front Environ Sci Eng* 2018; **12**: 1–10.

60	Ji L, Zhang Q, Fu X, *et al.* Feedback of airborne bacterial consortia to haze pollution with different PM2.5 levels in typical mountainous terrain of Jinan, China. *Sci Total Environ* 2019; **695**: 133912.

61	Karlsson E, Johansson AM, Ahlinder J, *et al.* Airborne microbial biodiversity and seasonality in Northern and Southern Sweden. *PeerJ* 2020; **2020**: 1–25.

62	Kaushik R, Balasubramanian R. Assessment of bacterial pathogens in fresh rainwater and airborne particulate matter using Real-Time PCR. *Atmos Environ* 2012; **46**: 131–9.

63	Li J, Cao J, Zhu Y, *et al.* Global Survey of Antibiotic Resistance Genes in Air. *Environ Sci Technol* 2018; : acs.est.8b02204.

64	Li H, Zhou X-Y, Yang X-R, Zhu Y-G, Hong Y-W, Su J-Q. Spatial and seasonal variation of the airborne microbiome in a rapidly developing city of China. *Sci Total Environ* 2019; **665**: 61–8.

65	Li H, Shan Y, Huang Y, *et al.* Bacterial community specification in PM2.5 in different seasons in Xinxiang, Central China. *Aerosol Air Qual Res* 2019; **19**: 1355–64.

66	Lu R, Li Y, Li W, *et al.* Bacterial community structure in atmospheric particulate matters of different sizes during the haze days in Xi’an, China. *Sci Total Environ* 2018; **637**–**638**: 244–52.

67	Mhuireach G, Betancourt-Román CM, Green JL, Johnson BR. Spatiotemporal controls on the urban aerobiome. *Front Ecol Evol* 2019; **7**. DOI:10.3389/fevo.2019.00043.

68	Mu F, Bai W, Li Y, Lu R, Qi Y, Xie W. Source identification of airborne bacteria in the mountainous area and the urban areas. *Atmos Res* 2020; **231**. https://dx.doi.org/10.1016/j.atmosres.2019.104676.

69	Najafi MS, Khoshakhllagh F, Zamanzadeh SM, Shirazi MH, Samadi M, Hajikhani S. Characteristics of TSP Loads during the Middle East Springtime Dust Storm (MESDS) in Western Iran. *Arab J Geosci* 2014; **7**: 5367–81.

70	Nanclares Castañeda DA, Zapata Sánchez CH, Silva-Bedoya LM, Montontoya Campuzáno OI, Moreno Herrera CX. Assessment of culturable bacteria associated with fine particulate matter collected in antioquia colombia-South America. *Rev Int Contam Ambient* 2020; **36**: 287–302.

71	Núñez A, Alcamí A, Amo de Paz G, *et al.* Temporal patterns of variability for prokaryotic and eukaryotic diversity in the urban air of Madrid (Spain). *Atmos Environ* 2019; published online Sept 8. https://dx.doi.org/10.1016/j.atmosenv.2019.116972.

72	Núñez A, Moreno DA. The Differential Vertical Distribution of the Airborne Biological Particles Reveals an Atmospheric Reservoir of Microbial Pathogens and Aeroallergens. *Microb Ecol* 2020; **80**: 322–33.

73	Pan Y, Pan X, Xiao H, Xiao H. Structural Characteristics and Functional Implications of PM2.5 Bacterial Communities During Fall in Beijing and Shanghai, China. *Front Microbiol* 2019; **10**: 1–11.

74	Raghav N, Mamta, Shrivastava JN, Satsangi GP, Kumar R. Enumeration and characterization of airborne microbial communities in an outdoor environment of the city of Taj, India. *Urban Clim* 2020; **32**. DOI:10.1016/j.uclim.2020.100596.

75	Rocha-Melogno L, Ginn O, Bailey ES, *et al.* Bioaerosol sampling optimization for community exposure assessment in cities with poor sanitation: A one health cross-sectional study. *Sci Total Environ* 2020; **738**. DOI:10.1016/j.scitotenv.2020.139495.

76	Rodriguez-Gomez C, Ramirez-Romero C, Cordoba F, *et al.* Characterization of culturable airborne microorganisms in the Yucatan Peninsula. *Atmos Environ* 2020; **223**: 117183.

77	Romano S, Becagli S, Lucarelli F, Rispoli G, Perrone MR. Airborne bacteria structure and chemical composition relationships in winter and spring PM10 samples over southeastern Italy. *Sci Total Environ* 2020; **730**: 138899.

78	Rosas I, Salinas E, Yela A, Calva E, Eslava C, Cravioto A. Escherichia coli in settled-dust and air samples collected in residential environments in Mexico City. *Appl Environ Microbiol* 1997; **63**: 4093–5.

79	Santos RA, Sau NJ, Certucha T, Almendariz FJ. Rapid detection of bacteria, Enterococcus faecalis, in airborne particles of Hermosillo, Sonora, México. *J Environ Biol* 2019; **40**: 619–25.

80	Santos-Burgoa C, Rosas I, Yela A. Occurrence of airborne enteric bacteria in Mexico city. *Aerobiologia (Bologna)* 1994; **10**: 39–45.

81	Serrano-Silva N, Calderon-Ezquerro MC. Metagenomic survey of bacterial diversity in the atmosphere of Mexico City using different sampling methods. *Environ Pollut* 2018; **235**: 20–9.

82	Shen F, Zheng Y, Niu M, *et al.* Characteristics of biological particulate matters at urban and rural sites in the North China Plain. *Environ Pollut* 2019; **253**: 569–77.

83	Soleimani Z, Sorooshian A, Goudarzi G, Maleki H, Marzouni MB. Impact of Middle Eastern dust storms on indoor and outdoor composition of bioaerosol. *Atmos Environ* 2016; **138**: 135–43.

84	Soto T, Lozano M, Vicente-Soler J, Cansado Vizoso J, Gacto Fernández MJ. Microbiological survey of the aerial contamination in urban areas of the city of Murcia, Spain. *An Biol* 2009; **31**: 7–13.

85	Stewart JD, Shakya KM, Bilinski T, Wilson JW, Ravi S, Choi CS. Variation of near surface atmosphere microbial communities at an urban and a suburban site in Philadelphia, PA, USA. *Sci Total Environ* 2020; **724**: 138353.

86	Sun Y, Xu S, Zheng D, Li J, Tian H, Wang Y. Effects of haze pollution on microbial community changes and correlation with chemical components in atmospheric particulate matter. *Sci Total Environ* 2018; **637**–**638**: 507–16.

87	Velez-Quinones MA. Study of Airborne Bacterial Populations in Urban Areas Using Phenotypic and Phylogenetic Characterization. ProQuest Diss. Theses. 2013. https://go.openathens.net/redirector/gatech.edu?url=https://search.proquest.com/docview/1438179215?accountid=11107.

88	Wei K, Zou Z, Zheng Y, *et al.* Ambient bioaerosol particle dynamics observed during haze and sunny days in Beijing. *Sci Total Environ* 2016; **550**: 751–9.

89	Wei M, Liu H, Chen J, *et al.* Effects of aerosol pollution on PM2.5-associated bacteria in typical inland and coastal cities of northern China during the winter heating season. *Environ Pollut* 2020; **262**: 114188.

90	Woo AC, Brar MS, Chan Y, *et al.* Temporal variation in airborne microbial populations and microbially-derived allergens in a tropical urban landscape. *Atmos Environ* 2013; **74**: 291–300.

91	Wu B, Meng K, Wei L, Cai Y, Chai T. Seasonal Fluctuations of Microbial Aerosol in Live Poultry Markets andthe Detection of Endotoxin. *Front Microbiol* 2017; **8**. DOI:10.3389/fmicb.2017.00551.

92	Xu A, Chen X, Lang X, Xia Y, Song Z. Seasonal variability in bacterial and fungal diversity and community composition of the near-surface atmosphere in coastal megacity. *Aerobiologia (Bologna)* 2017; **33**: 555–75.

93	Xu C, Wei M, Chen J, *et al.* Bacterial characterization in ambient submicron particles during severehaze episodes at Ji’nan, China. *Sci Total Environ* 2017; **580**: 188–96.

94	Yan D, Tao Z, Yu L-Y, Su; J, Yu L-Y. Structural Variation in the Bacterial Community Associated with Particulate Matter in Beijing, China During Hazy and Nonhazy Days. *Appl Environ Microbiol* 2018; **84**: 1–13.

95	Yassin MF, Almouqatea S. Assessment of airborne bacteria and fungi in an out door environment. *Int J fo Environ Sci Technol* 2010; **7**: 535–44.

96	Yooseph S, Andrews-Pfannkoch C, Tenney A, *et al.* A metagenomic framework for the study of airborne microbial communities. *PLoS One* 2013; **8**. DOI:10.1371/journal.pone.0081862.

97	Zhang S, Du R, Chen H, Ren W, Du P. Seasonal variation of microbial activity and pathogenic bacteria under non-serious pollution levels in Beijing. *Aerosol Air Qual Res* 2019; **19**: 1798–807.

98	Zhen Q, Deng Y, Wang Y, *et al.* Meteorological factors had more impact on airborne bacterial communities than air pollutants. *Sci Total Environ* 2017; **601**–**602**: 703–12.

99	Zhou H, Wang X, Li Z, Kuang Y, Mao D, Luo Y. Occurrence and Distribution of Urban Dust-Associated Bacterial Antibiotic Resistance in Northern China. *Environ Sci Technol Lett* 2018; **5**: 50–5.
